# Fine-Grained Forecasting of COVID-19 Trends at the County Level in the United States

**DOI:** 10.1101/2024.01.13.24301248

**Authors:** Tzu-Hsi Song, Leonardo Clemente, Xiang Pan, Junbong Jang, Mauricio Santillana, Kwonmoo Lee

**Affiliations:** Vascular Biology Program and Department of Surgery, Boston Children’s Hospital, Harvard Medical School, Boston, MA 02115, USA; Department of Physics and Department of Electrical and Computer Engineering, Northeastern University, Boston, MA 02115, USA

## Abstract

The novel coronavirus (COVID-19) pandemic, first identified in Wuhan China in December 2019, has profoundly impacted various aspects of daily life, society, healthcare systems, and global health policies. There have been more than half a billion human infections and more than 6 million deaths globally attributable to COVID-19. Although treatments and vaccines to protect against COVID-19 are now available, people continue being hospitalized and dying due to COVID-19 infections. Real-time surveillance of population-level infections, hospitalizations, and deaths has helped public health officials better allocate healthcare resources and deploy mitigation strategies. However, producing reliable, real-time, short-term disease activity forecasts (one or two weeks into the future) remains a practical challenge. The recent emergence of robust time-series forecasting methodologies based on deep learning approaches has led to clear improvements in multiple research fields. We propose a recurrent neural network model named Fine-Grained Infection Forecast Network (FIGI-Net), which utilizes a stacked bidirectional LSTM structure designed to leverage fine-grained county-level data, to produce daily forecasts of COVID-19 infection trends up to two weeks in advance. We show that FIGI-Net improves existing COVID-19 forecasting approaches and delivers accurate county-level COVID-19 disease estimates. Specifically, FIGI-Net is capable of anticipating upcoming sudden changes in disease trends such as the onset of a new outbreak or the peak of an ongoing outbreak, a skill that multiple existing state-of-the-art models fail to achieve. This improved performance is observed across locations and periods. Our enhanced forecasting methodologies may help protect human populations against future disease outbreaks.

## 1 Introduction

Since 2019, the SARS-CoV-2 coronavirus has spread in human populations and causing a disease called COVID-19. Its rapid spread, from several countries to the global stage, prompted the World Health Organization (WHO) to declare it a global pandemic in early 2020 [13]. With over half a billion infections and more than 6 million deaths recorded worldwide [15], COVID-19 has significantly reshaped society, national healthcare systems, and the global economy. Due to its fast mutation rate, new outbreaks of COVID-19 have continued to emerge within a span of a few months, making forecasting the trends of these COVID-19 waves a crucial task that helps public health officials to regulate public healthcare policies and efficiently manage medical resources to prevent and control future outbreaks [17].

Epidemic forecasting in this context has presented a formidable challenge, driving the development of numerous methods to address this complexity. With the advent of the COVID-19 pandemic, the need for efficient preventive measures and accurate forecasting models tailored to this emerging infectious disease became increasingly critical. Within the field of epidemiology, Susceptible–Infected–Removed (SIR) models and their variations are extensively utilized to assess the spread of infections [27, 23]. These epidemiological models allow us to deduce crucial parameters such as infection and death rates, enabling the forecast of disease transmission trends. Several COVID-19 studies have demonstrated practical insights and forecasts by employing SIR-like models in various regions worldwide [33, 28, 34, 14, 38, 2, 40, 4, 21, 9, 46, 54, 10, 29]. However, spatial and temporal heterogeneity known to exist across locations, such as socioeconomic and demographic factors, the diverse implementation of local mitigation policies, and the emergence of new variants, are often very challenging to incorporate and appropriately update in these models. As a result, these methods often fall short in capturing essential local heterogeneities in infection dynamics [12, 48].

In contrast, data-driven methods may excel in this context by implicitly incorporating the influence of diverse local policies, new variants, demography, and socioeconomic factors, by learning directly from the reported data. Notably, the United States displays highly diverse vaccination rates, adding complexity to the accurate characterization and prediction of disease spread. Hence, alternative data-driven models could offer complementary insights into disease dynamics [52].

Data-driven machine learning models have been extensively applied in disease forecasting [8, 49, 50]. For instance, Sujath *et al.* conducted a comparison of linear regression, multilayer perceptron (MLP), and vector autoregression, with the MLP model demonstrating superior performance [51]. Ardabili *et al.* integrated genetic algorithms with SIR and SEIR models to predict outbreak trends [3]. Hernandez *et al.* utilized an Auto-regressive Integrated Moving Average (ARIMA) [20] model with polynomial functions for global COVID infection trend predictions, using cumulative datasets [22]. Additionally, Lu *et al.* compared and integrated various approaches, combining statistical linear and nonlinear models to estimate the cumulative number of weekly confirmed cases [36]. Many of these models rely on assumptions about the real-time availability of reliable data, a stable non-evolving pathogen, and/or population-level adherence to specific behaviors during epidemic outbreaks. However, the dynamics of COVID-19 have been in constant flux, influenced by factors such as virus characteristics and heterogeneous population adherence to mitigation policies such as recommendations to stay at home or mask wearing [30]. Consequently, COVID-19 forecasting has become an exceptionally challenging endeavor. Deep learning’s feature-learning ability holds promise for addressing challenges stemming from the local heterogeneity of infection dynamics, especially after enough observations to train models have been recorded. Several studies have utilized deep learning methods to predict COVID-19 data, incorporating related time series models such as Long Short-Term Memory (LSTM) networks, Convolutional Neural Networks (CNN), and hybrid approaches that combine deep learning with traditional methods. Rodriguez *et al.* utilized a feed-forward network to generate short-term forecasts of COVID-19, demonstrating the responsive capacity of their models to adapt to sparse data situations. Bandyopadhyay *et al.* used LSTM with a gate circulation network to estimate COVID-19 cases [16], while Huang *et al.* employed CNNs to predict cumulative COVID-19 deaths [32]. Ruifang *et al.* combined LSTM with a Markov method for national cumulative COVID-19 predictions [37]. ArunKumar *et al.* compared the performance of deep learning models (GRU and LSTM) with traditional models like ARIMA and SARIMA, concluding that deep learning models were better suited for non-linear datasets [5]. Additionally, Transformer models, known for their proficiency in natural language processing, have been utilized in COVID-19 forecasting. Soumyanil *et al.* developed a graph transformer network with synchronous temporal and spatial information [6], while Kapoor *et al.* used a similar approach to predict COVID-19 confirmed cases [26]. However, these models were based on limited national or state-level datasets, where the local heterogeneity of COVID-19 infections was averaged out. Consequently, despite the impressive capabilities of deep learning, their forecasting results were restricted, especially during the rapidly changing outbreak of the COVID-19 variants such as Omicron.

In response to these challenges, we present a novel deep learning approach that harnesses extended time series data covering the entire span of the COVID-19 pandemic in the United States, including the Omicron wave. Our model, built on the foundation of Long Short-Term Memory (LSTM) networks and incorporating stacked bidirectional components, capitalizes on temporal relation awareness to adeptly manage abrupt shifts in infection dynamics. This ensures precise short-term predictions, vital for shaping future epidemic prevention and control strategies. Our method, named FIGI-Net (FineGrained Infection Forecast Network), delves into fine-grained time series of COVID-19 infection data at the county level in the U.S.. This approach leverages LSTM capabilities with a stacked bidirectional component, enhancing the model’s capacity to discern diverse global infection trends. Subsequently, upon identifying clusters of counties with similar COVID-19 temporal patterns, we applied transfer learning from the global biLSTM model trained on segmented historical daily confirmed cases of U.S. counties, and trained specific cluster-based biLSTM models. This cluster transfer learning concept preserves the model’s inherent capabilities [47, 55, 44, 25, 45] while empowering it to swiftly adapt to short-term trend changes locally, refining forecasting accuracy. Using our proposed FIGI-Net, we conducted predictions for future COVID-19 infection cases, encompassing both short-term and midterm forecasts spanning from the next day to the next 15 days. The key contributions of our framework are summarized as follows:

- We provide a practical approach to determine the minimum amount of observations –length of the initial training time period – needed for deep learning-based machine learning models to deliver reliable daily disease activity forecasts in the context of a novel and emerging disease outbreak.
- We introduce FIGI-Net (Fine-Grained Infection Forecast Network), a deep learning pipeline that leverages COVID-19 infection time series data of U.S. counties, and is capable of forecasting COVID-19 confirmed cases up to 15 days ahead. Harnessing bidirectional temporal feature learning and transfer learning techniques, our model was trained with infectious clusters of COVID-19 temporal data. The structure of the proposed framework is detailed in Figure 1 and more details are presented in Section 4.C.
- Given the explicit need to produce a model responsive and adaptive to dynamic changes of disease transmission due to a diverse set of factors – changes in human behavior, availability of vaccines, and testing capabilities, our model is dynamically re-trained on a moving-window that only uses the most recent trends as input. This time window is chosen prior to evaluating our model’s forecasts in a strictly out-of-sample fashion.
- Focusing on U.S. county-level COVID-19 time series data, we show FIGI-Net’s efficiency and accuracy in forecasting sharp changes in COVID-19 activity, both in short-term (up to 1 day) and mid-term (up to 2 weeks) forecasting scenarios.
- Furthermore, our methodology is extended to national and state-level forecasts using weekly time periods. The results highlight the superiority of our FIGI-Net, showcasing more than a 40% error reduction during critical time periods compared to other state-of-the-art models designed for COVID-19 infection forecasting tasks.

**Figure 1:**
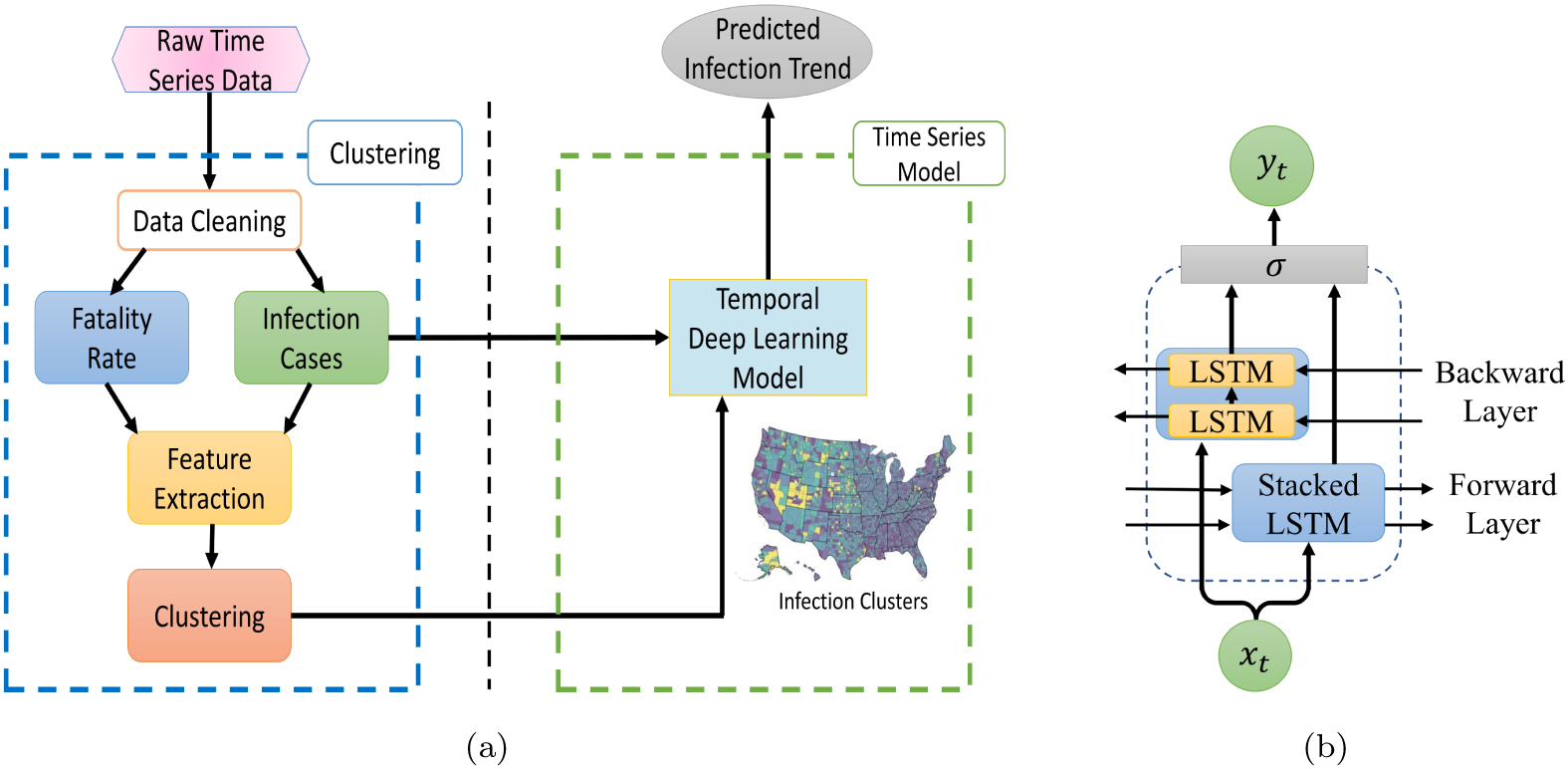
Our proposed model, FIGI-Net, architecture for COVID-19 infection prediction. (a) A diagram that visualizes our methodology’s framework, consisting of a clustering and a time series deep learning component. In the clustering section, auto-correlation and cross-correlation were used to extract similar features from the pre-cleaning infection cases and fatality rate data. Then, we applied a clustering technique to the extracted features to identify similar COVID-19 dynamics. The temporal deep learning model utilized the given clusters along with the infection case data to predict the trend of COVID-19. (b) The architecture of the proposed bidirectional stacked LSTM model. This model learns the feature dependencies of input sequential data in both forward and backward directions and can effectively deal with short-term state changes. The *σ* function represents the merging function.

## Results

We implemented a collection of machine-learning based models to generate out-of-sample predictions for the number of COVID-19 confirmed cases, as reported by the Centers for Disease Control and Prevention (CDC), for the time period between October 18^th^, 2020 and April 15^th^, 2022. These models included the model we propose: FIGI-Net, as well as Autoregressive statistical models, a collection of neural network based models (GRU, LSTM), a stacked bidirectional LSTM (biLSTM), a set of LSTM-based models incorporating temporal clustering (TC-LSTM and TC-biLSTM), and a ”naive” (Persistence) model to be used as a baseline.

We used our models to retrospectively produce daily forecasts for multiple time horizons, *h*, ranging from *h* = 1, 2*…,* 15 days ahead. Visual representations of our predictions, alongside the actual observed COVID-19 confirmed cases, are presented in Figures 3, 4, and 5 for county, state and national levels. We evaluated our model’s performance by comparing our out-of-sample predictions with subsequently observed reported data in each time horizon *h* using multiple error metrics that include: the root mean square error (RMSE), relative RMSE (RRMSE), and mean absolute percentage error (MAPE) metrics, as detailed in Section 4.D. In addition, we compared the prediction performance of our model with a diverse set of statistical and machine learning models, including state-of-the-art forecasting methodologies reported by the CDC (a comprehensive list of these models is provided in Section 2.G). Finally, we assessed our model’s ability to anticipate the onset of multiple outbreaks during our studied time periods.

### Determining the Length of Moving Time Windows for Training

Given that neural network based models typically need a large amount of data to be trained [31, 56], we first investigated the minimum amount of data that would be necessary for our model to produce robust and reliable forecasts. We investigated the length of the moving window for training necessary to yield forecasts responsive to changes in disease dynamics due to changes in human behavior over time, vaccination adherence and vaccine availability, different transmission intensities for different variants of concern (e.g. omicron), among other factors. Using observations from April 18^th^, 2020 to October 17^th^, 2020 we identified a time window size of 75 days to be a good compromise between reliable forecasting performance (shown in Figure 2) and a short enough time period that would allow the model to continuously learn new transmission patterns. For more details, please refer to the Section Discussion.

**Figure 2:**
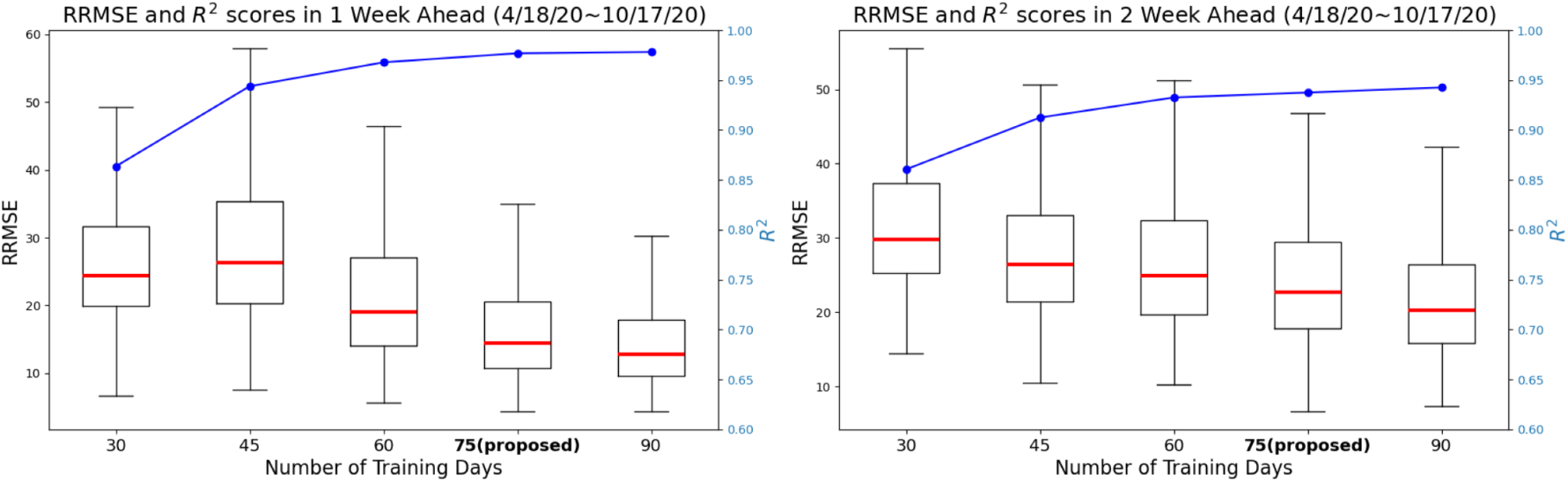
Comparison of training day length identification for the proposed model in 1 and 2 weekly horizons. The figure shows the RRMSE values and *R*^2^ scores (represented by the blue line) achieved by the proposed model using different training day lengths from first 6-month dataset (from April to Oct, 2020). Here For the 1 weekly horizon, 75-day and 90-day training lengths result in lower RRMSE prediction errors, with a median value below 20 and an approximate *R*^2^ score of 0.95. Moreover, increasing the training day length beyond 75 days leads to a 26.3% reduction in RRMSE for 2 weekly ahead prediction but does not further improve performance with longer training periods. These findings highlight the significant impact of training day length on short-term infection forecasting.

### FIGI-Net Forecasting Performance at County Level

For each forecasting horizon *h*, we computed FIGI-Net’s prediction error metrics (RMSE and RRMSE) across all counties, over the time period: 10/18/2020 - 4/15/2022. Table 1 shows the prediction error values (and percentage of error reduction with respect to the Persistence model) of all the models for 1-day, 7-day and 14-day horizons. Based on the experiments regarding training length influence, we evaluated all comparative models using an optimal training length of 75 days in the following eighteen months of data. Our results demonstrate that FIGI-Net model has the greatest error reduction across all horizons (90%, 83%, and 45% RMSE reduction, correspondingly), Followed by the biLSTM model (85% in horizon 1-day, 75% in horizon 7-day, and 44% in horizon 14-day RMSE reduction). To assess the significance of these error reductions, we performed the two-sided Wilcoxon rank sum test [42] over the entire outcomes of tasks.

**Table 1:** Performance metrics with error reduction for the 1-day, 7-day and 14-day ahead tasks at the county level.

| Model | RMSE |  |  | RRMSE(%) |  |  | Error Reduction |  |  |
| --- | --- | --- | --- | --- | --- | --- | --- | --- | --- |
|  | 1<sup>st</sup> day | 7<sup>th</sup> day | 14<sup>th</sup> day | 1<sup>st</sup> day | 7<sup>th</sup> day | 14<sup>th</sup> day | 1<sup>st</sup> day | 7<sup>th</sup> day | 14<sup>th</sup> day |
| Persistence | 45.11±176.33 | 29.71±133.62 | 36.56±152.9 | 83.13±111.19 | 50.48±48.18 | 65.48±69.41 | — | — | — |
| Autoregressive | 44.43±209.3 | 49.15±9.68x10<sup>4</sup> | 65.91±3.17x10<sup>9</sup> | 81.57±95.71 | 83.87±3.18x10<sup>4</sup> | 103.16±8.72x10<sup>8</sup> | 1.01±1.13 | 1.5±702.98 | 1.54±1.12x10<sup>7</sup> |
| GRU | 18.62±101.65 | 19.23±106.35 | 23.75±118.99 | 33.26±24.04 | 34.45±25.11 | 43.61±25.27 | 0.49±0.3 | 0.68±0.4 | 0.7±0.33 |
| LSTM | 18.44±91.71 | 18.01±104.53 | 22.83±118.66 | 31.29±24.09 | 33.37±25.08 | 42.04±24.9 | 0.46±0.3 | 0.64±0.4 | 0.7±0.33 |
| TC-LSTM | 19.03±105.94 | 19.22±112.28 | 23.56±123.17 | 33.19±23.1 | 35.09±26.23 | 43.84±25.97 | 0.49±0.3 | 0.7±0.4 | 0.72±0.34 |
| TC-biLSTM | 10.47±59.91 | 12.96±80.49 | 23.66±115.85 | 17.08±20.25 | 23.03±30.25 | 44.33±37.06 | 0.25±0.24 | 0.43±0.51 | 0.73±0.4 |
| biLSTM | 6.53±34.9 | 8.29±62.98 | 20.51±98.42 | 9.83±18.41 | 14.28±34.5 | 40.01±34.98 | 0.15±0.22 | 0.25±0.54 | 0.66±0.4 |
| FIGI-Net | **4.34±26.44** | **5.98±61.22** | **19.79±100.08** | **6.98±17.73** | **9.92±37.31** | **39.73±34.99** | **0.1±0.22** | **0.17±0.57** | **0.65±0.42** |

In Figure 3.A, we visualize the forecasting ability of each model for both the 1-day and 7-day ahead task, including classic models such as Persistence (a naive rule stating *y_t_*_+1_ = *y_t_*) and Autoregressive models(AR) [7], and deep learning-based models such as GRU or LSTM architectures. We used the median RMSE score for each model as a means to order them in decreasing order (leftmost model with the worst performance, and rightmost with the best). Our first observation is that FIGI-Net scored the lowest median RMSE and relative RMSE scores across all models for both the 1-day ahead and the 7-day ahead prediction task (approximately 6.98% at 1-day ahead, and 9.92% at 7-day ahead), followed by deep learning models with bidirectional components (TC-biLSTM with a reduction of 17.08% and 23.03% and biLSTM with a reduction of 9.83% and 14.28%, respectively). Generally speaking, all network based models improved over Persistence (with an error of 83.13% and 50.48%) and the Autoreggressive models (81.57% and 83.87%). For a detailed description of the performance of each model, please refer to Table 1.

**Figure 3:**
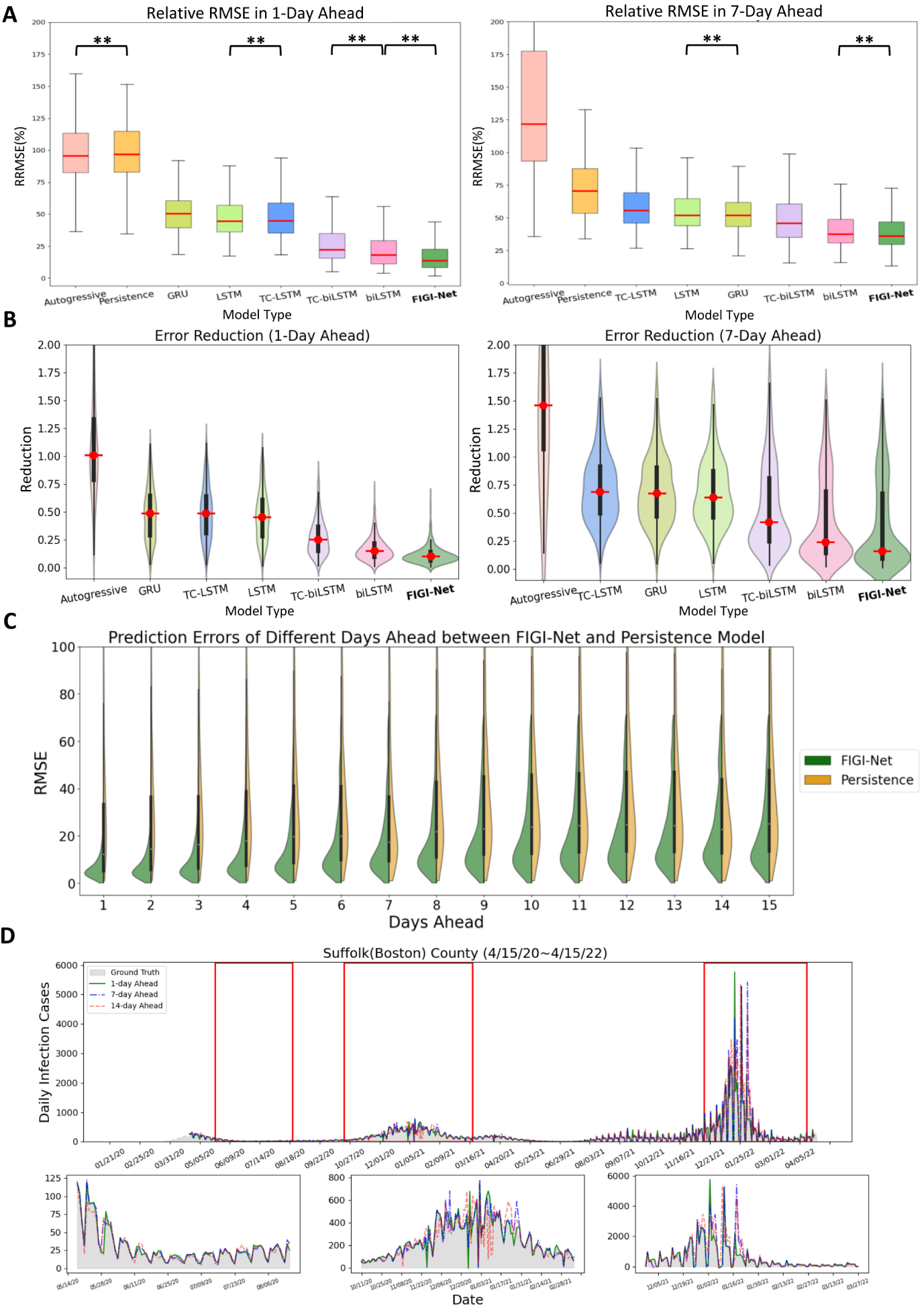
A summary of the comparative performance of FIGI-Net at the county level, for the 1-day and 7-day ahead horizon tasks from Oct. 18^th^, 20 to Apr. 15^th^, 22. (A) Performance of each model over all 3143 counties presented as a box plot. The median is highlighted in red along with the 5^th^ and 95^th^ percentile whiskers. The models are ordered in decreasing order, with the most accurate model (lowest RRMSE) appearing on the rightmost side. Notably, FIGI-Net exhibits the lowest RRMSE compared to other models for both tasks, as confirmed by the two-sided Wilcoxon rank sum test. (B) Error Reduction between each model with Persistence model. Compared to other models, FIGI-Net provides the fewest erroneous forecasting results. (C) Performance comparison of FIGI-Net against Persistence (our baseline model).Comparison is shown as a set of violin plots across the different time horizons. Our model consistently displays a narrower distribution of prediction errors (RMSE) compared to the Persistence model. (D) shows Daily prediction results of our model in 1-day, 7-day and 14-day horizons in Suffolk county. The example demonstrates our model’s ability to provide highly accurate predictions for diverse locations and various time periods. *∗∗*: p-value ¡ 0.001

Figure 3.B focuses on the performance of FIGI-Net against Persistence. For each time horizon, we generated a violin plot to visualize the RMSE scores of FIGI-Net (in blue) and Persistence (in orange) across all counties. Our results show that FIGI-Net has a higher concentration of scores between the 0-20 range across all time-horizons, in comparison to Persistence, where the scores of the orange distribution are widely spread. The main error difference between FIGI-Net and Persistence occur at the horizon 1, with a mean RMSE score of 9.97, in comparison to 79 from Persistence (a 89% reduction).

Figure 3.C showcases a visualization of the forecasts of our model for the county of Suffolk, Massachusetts, for the 1-day, 7-day and 14-day ahead tasks. Additionally to the full time period of the experiment, three periods from May to August 2020, October 2020 to February 2021, and December 2021 to March 2022, are also displayed. FIGI-Net accurately forecasted the daily infection trends in 1-day and 7-day ahead horizons in diverse counties, even when these counties exhibited contrasting infection trends, as shown for the country of Suffolk during the initial outbreak stages. However, we observed that the 14-day ahead forecasting trend yielded larger errors, particularly in cases where the infection numbers fluctuated significantly.

### Forecasting Performance at State Level

Similar to our county level experiment, we compared the performance of FIGI-Net against state-of-theart models for the state-level geographical resolution. For FIGI-Net, which is a model that leverages high volumes county-level activity as part of its design, we decided to aggregate the county level forecasts into state level (details regarding the performance of FIGI-Net, trained on state-level data, can be found in our supplementary materials Table S1). For the rest of the models, we trained them using State level data to obtain state-level outcomes. Then, we computed the RRMSE for each state across 1-day, 7-day, and 14-day ahead horizons, as shown in Figure 5.A. Our results, also shown in Figure 4 and summarized in Table 2, show that FIGI-Net was able to score 1149.66 1850.66 and 1935.36 3458.21 in terms of RMSE for the 7-day and 14-day horizon, correspondingly. On the other hand, the score of the Persistence model was 2571.9 5409.64 and 3330.82 6072.91 (resulting in an error reduction of 54%, and 39% in each case). Next to FIGI-Net, we can see the biLSTM (with a score of 1198.43 1806.38 and 1940.18 3216.91) and TC-biLSTM models (1338.63 2324.23 and 2225.14 4116.09 in RMSE).

**Figure 4:**
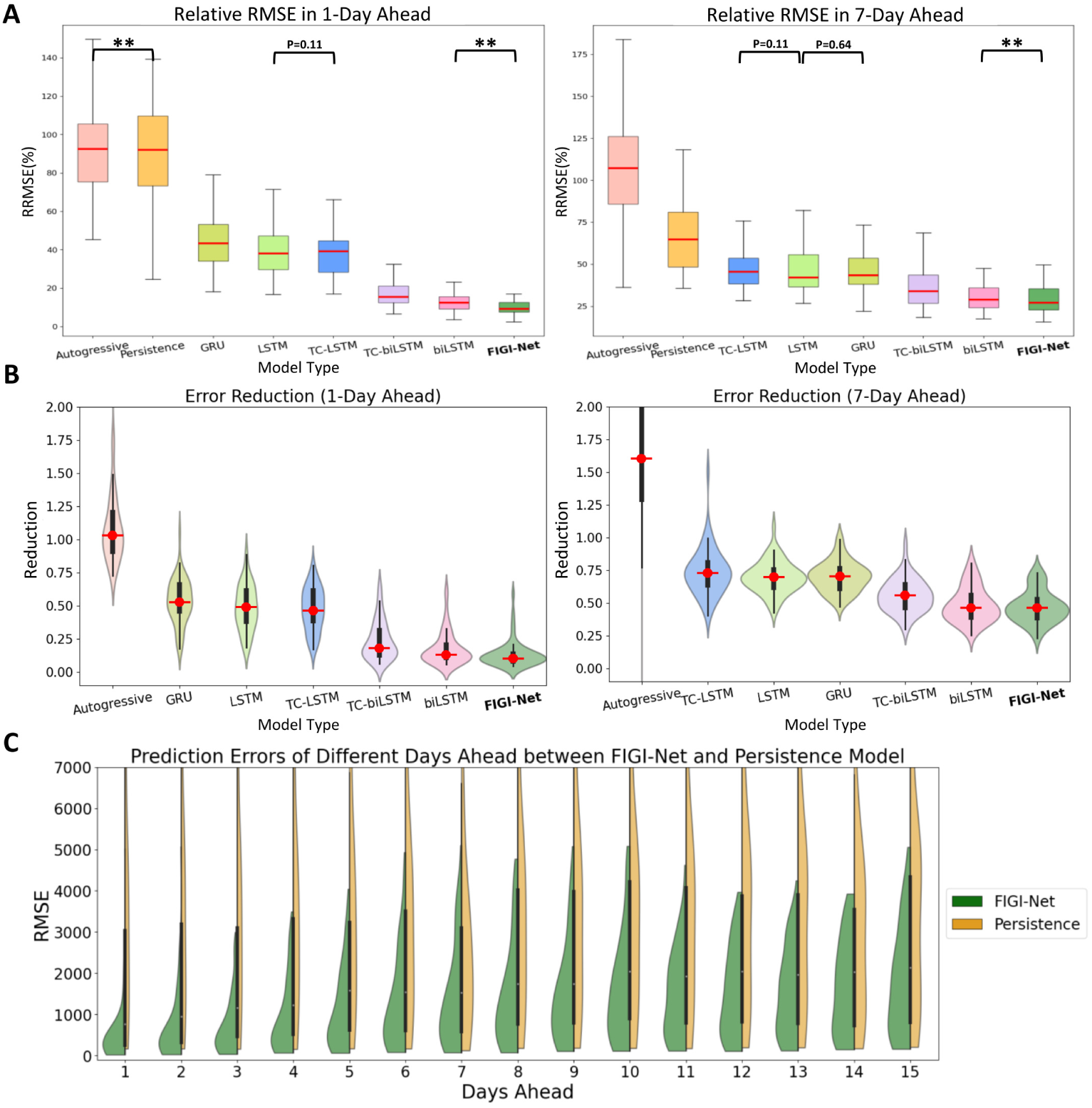
A comparative analysis of FIGI-Net’s performance at the state level. (A) At state level, FIGINet still exhibits the lowest RRMSE compared to other models for the 1-day and 7-day ahead horizon tasks. (B) Error Reduction at state level displays that FIGI-Net provides lower forecasting errors than other models and reduced errors by at least 53% compared to the Persistence model. (C) Performance comparison of FIGI-Net against Persistence at state level. This comparison also represents that our model consistently displays a much narrower distribution of prediction errors (RMSE) and provides much lower forecasting errors during the first four days. *∗∗*: p-value ¡ 0.001

**Figure 5:**
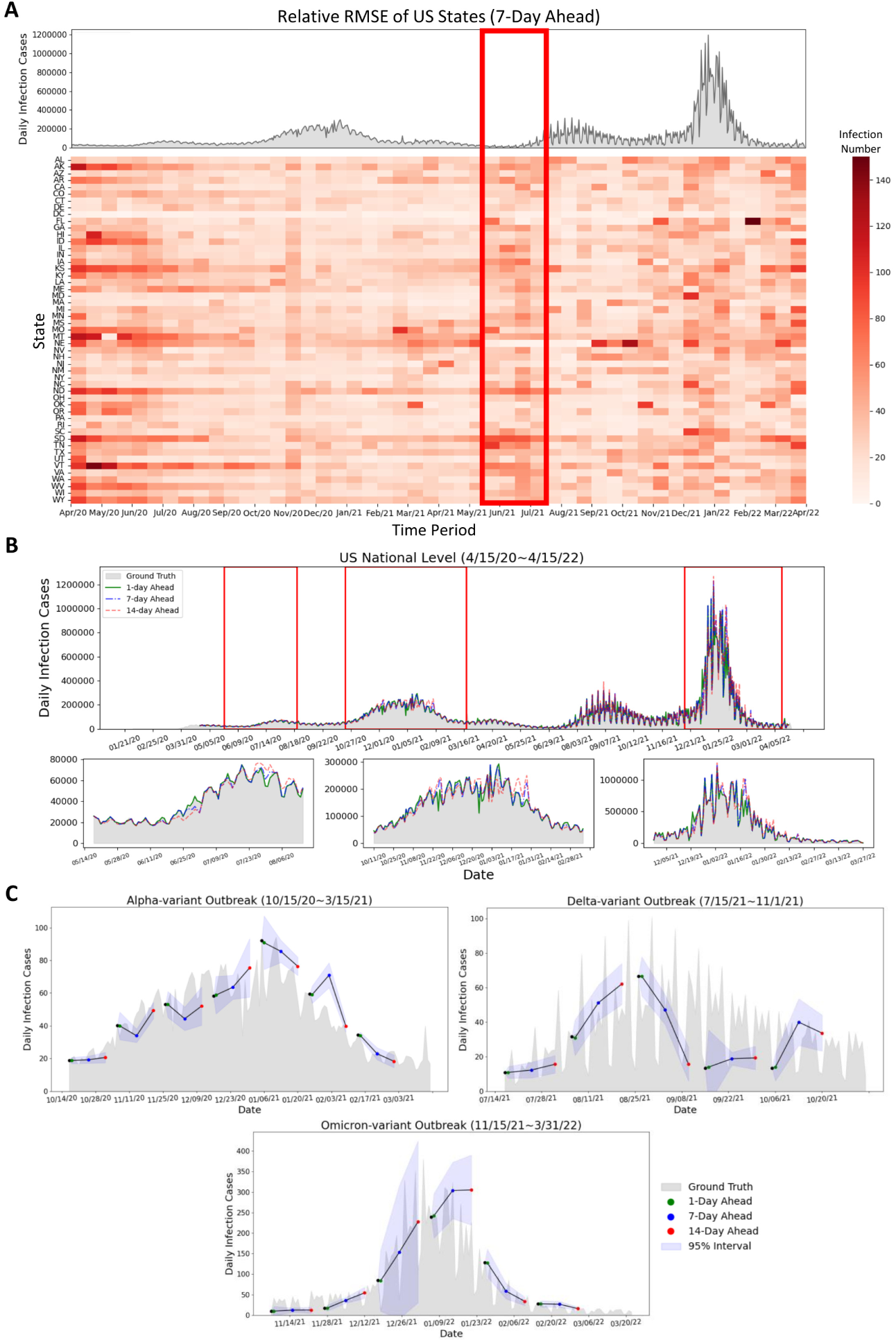
Summary of forecasting results at the state and national level. (A) Relative RMSE performance among US states in the 7-day horizon at the state level is determined by the average relative RMSE of the last 7 days of each time period, compared to the national reported infection trend. The relative RMSE increased during the early period (April 2020 to May 2020) of the first upward trend, the time period (June 2021 to July 2021) before the Delta outbreak, and the increasing period (December 2021 to January 2022) of the Omicron COVID-19 outbreak. Missouri, Montana, and Nebraska have large relative RMSE values during March 2021 to May 2021. We can observe that the RRMSE errors often increase before the early stage of the next outbreaks or when the infection trend rapidly increases (red rectangle) (B) Daily prediction infection trends during different days ahead at the national level. It can be observed that the daily predicted infection trends at 1-day and 7-day ahead horizons show similarity to the reported data, while the 14-day ahead trend exhibits some fluctuations. (C) Daily predicted infection trends of the proposed model during Alpha-variant, Delta-variant, and Omicron-variant outbreaks with different days ahead predictions. The proposed FIGI-Net model can provide a curve of predicted trends matching the observed report. The range of margin of error became larger when the trend of Omicron-variant outbreak increased.

**Table 2:** Performance metrics with error reduction for the 1-day, 7-day and 14-day ahead tasks at the state Level.

| Model | RMSE |  |  | RRMSE(%) |  |  | Error Reduction |  |  |
| --- | --- | --- | --- | --- | --- | --- | --- | --- | --- |
|  | 1<sup>st</sup> day | 7<sup>th</sup> day | 14<sup>th</sup> day | 1<sup>st</sup> day | 7<sup>th</sup> day | 14<sup>th</sup> day | 1<sup>st</sup> day | 7<sup>th</sup> day | 14<sup>th</sup> day |
| Persistence | 3563.29±7636.32 | 2571.9±5409.64 | 3330.82±6072.91 | 92.03±29.74 | 64.74±21.38 | 79.85±15.76 | — | — | — |
| Autoregressive | 3856.01±8189.29 | 4023.77±1.26x10<sup>6</sup> | 5932.29±3.29x10<sup>10</sup> | 92.47±28.45 | 107.09±1.61x10<sup>4</sup> | 160.23±4.82x10<sup>8</sup> | 1.03±0.32 | 1.62±217.8 | 2.23±5.72x10<sup>6</sup> |
| GRU | 1411.09±3827.44 | 1460.49±3947.68 | 1991.35±4503.38 | 43.29±16.38 | 43.51±15.36 | 56.06±14.06 | 0.53±0.19 | 0.71±0.18 | 0.67±0.12 |
| LSTM | 1332.83±3342.72 | 1782.58±3796.64 | 2028.5±4462.46 | 38.02±16.33 | 42.08±15.37 | 53±15.53 | 0.49±0.19 | 0.7±0.17 | 0.67±0.13 |
| TC-LSTM | 1311.08±4059.58 | 1947.45±4131.61 | 2021.77±4686.08 | 39.06±16.69 | 45.5±14.64 | 52.98±13.81 | 0.47±0.17 | 0.73±0.21 | 0.68±0.11 |
| TC-biLSTM | 550.36±1557.85 | 1338.63±2324.23 | 2225.14±4116.09 | 15.36±17.16 | 33.9±14.38 | 54.94±15.75 | 0.18±0.17 | 0.56±0.15 | 0.67±0.16 |
| biLSTM | 371±1047.57 | 1189.43±1806.38 | 1940.18±3216.91 | 12.44±15.04 | 28.9±14.01 | 49.46±17.18 | 0.13±0.15 | 0.46±0.18 | 0.63±0.22 |
| FIGI-Net | **290.31±908.81** | **1149.66±1850.66** | **1935.36±3458.21** | **9.17±13.59** | **26.94±14.48** | **49.12±16.47** | **0.11±0.13** | **0.46±0.15** | **0.61±0.21** |

During the early stages of the pandemic, we observe the RRMSE values of FIGI-Net were notably higher. For instance, based on Figure 5.A, Missouri, Montana, and Nevada displayed larger prediction errors between March to May 2021, as shown from the deep red color in the center of the RRMSE matrix. During this period, these states experienced a significant spike in COVID-19 activity, deviating from both previous months and the overall national trend. Alternatively, the red rectangle in Figure 5.A illustrates that the RRMSE errors often increase before the outbreaks or when the infection trend rapidly increases. This observation suggests that the proposed FIGI-Net model may provide early prediction outcomes for outbreaks. Similar trends are also evident in other day-ahead forecasting instances (shown in Figure S4).

### National Forecasting Performance

Figure 5.B represents national level COVID-19 official reports contrasted with our model predictions across three different horizons. Similar to the observations in Figure 3.C, our predictions were highly accurate at the 1-day and 7-day ahead horizons, with the RRMSE of 7.02% and 15.47%, respectively, compared to the national official reports. However, at the 14-day horizon, the discrepancies between predicted values and ground truth grew during high infection periods, resulting in an RRMSE of 27.94%. Table 3 shows the performance between FIGI-Net and other models at national level. Compared to our benchmark models, FIGI-Net improved forecast capacity can lead to a 86.5% reduction at 1-day horizon, a 60.98% reduction at 7-day horizon, and a 53.8% reduction at 14-day horizon in RRMSE score.

**Table 3:**
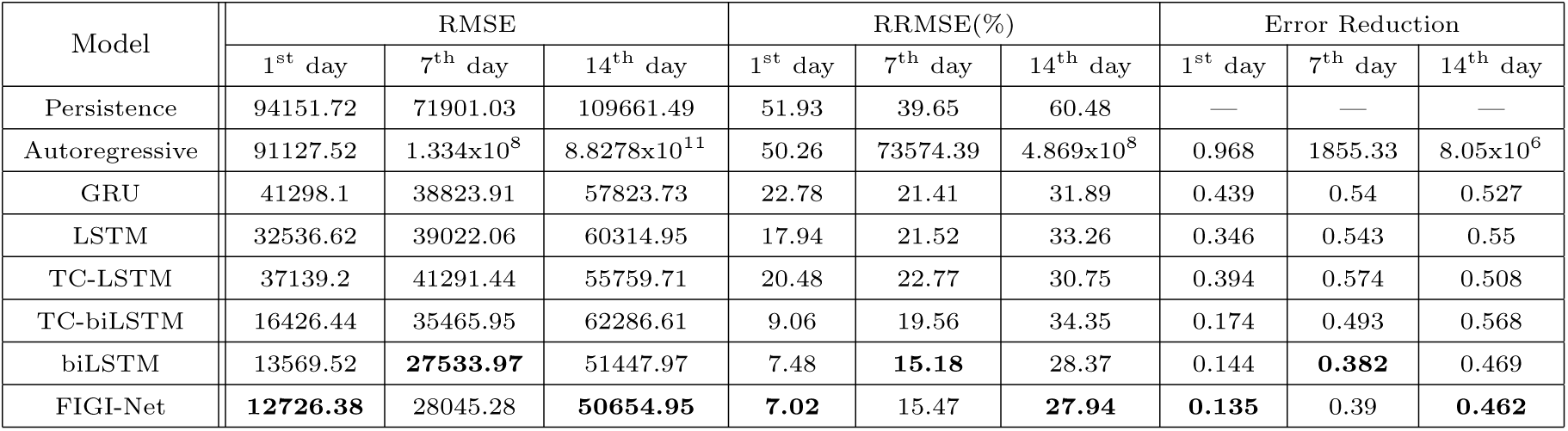
Performance metrics with error reduction for the 1-day, 7-day and 14-day ahead tasks at the national Level.

| Model | RMSE |  |  | RRMSE(%) |  |  | Error Reduction |  |  |
| --- | --- | --- | --- | --- | --- | --- | --- | --- | --- |
|  | 1<sup>st</sup> day | 7<sup>th</sup> day | 14<sup>th</sup> day | 1<sup>st</sup> day | 7<sup>th</sup> day | 14<sup>th</sup> day | 1<sup>st</sup> day | 7<sup>th</sup> day | 14<sup>th</sup> day |
| Persistence | 94151.72 | 71901.03 | 109661.49 | 51.93 | 39.65 | 60.48 | — | — | — |
| Autoregressive | 91127.52 | 1.334x10<sup>8</sup> | 8.8278x10<sup>11</sup> | 50.26 | 73574.39 | 4.869x10<sup>8</sup> | 0.968 | 1855.33 | 8.05x10<sup>6</sup> |
| GRU | 41298.1 | 38823.91 | 57823.73 | 22.78 | 21.41 | 31.89 | 0.439 | 0.54 | 0.527 |
| LSTM | 32536.62 | 39022.06 | 60314.95 | 17.94 | 21.52 | 33.26 | 0.346 | 0.543 | 0.55 |
| TC-LSTM | 37139.2 | 41291.44 | 55759.71 | 20.48 | 22.77 | 30.75 | 0.394 | 0.574 | 0.508 |
| TC-biLSTM | 16426.44 | 35465.95 | 62286.61 | 9.06 | 19.56 | 34.35 | 0.174 | 0.493 | 0.568 |
| biLSTM | 13569.52 | **27533.97** | 51447.97 | 7.48 | **15.18** | 28.37 | 0.144 | **0.382** | 0.469 |
| FIGI-Net | **12726.38** | 28045.28 | **50654.95** | **7.02** | 15.47 | **27.94** | **0.135** | 0.39 | **0.462** |

Additionally, we analyzed the daily performance of FIGI-Net at the national level across three main outbreak waves: the Alpha-variant wave (October 15^th^, 2020 to March 15^th^, 2021), Delta-variant wave (July 15^th^, 2021 to November 1^st^, 2021), and Omicron-variant outbreak wave (November 15^th^, 2021 to March 31^st^, 2022). By aggregating our county-level forecasts, we depicted the national prediction trends during these waves (Figure 5.C). Our model demonstrated an accurate prediction direction, maintaining an average error range of 12.17 infection cases at a confidence level of 95% during the Alpha-variant and Delta-variant waves. During the Omicron-variant wave, the forecasting trend exhibited a wide range of confidence intervals. However, our model still successfully predicted trends up to the 14-day horizon. These above outcomes underscore the robustness of our FIGI-Net model in addressing substantial variations in infection numbers.

### Geographical Distribution of COVID-19 Infection Predictions of US Counties

In this section, we conducted a geo-spatial analysis with the objective to identify possible geographical patterns in the performance of FIGI-Net. Figure 6 illustrates various geographical maps of US counties. The first column of Sections a, b, and c presents the average number of cases occurring for 3 distinct 1-week periods: the first halves of August 2020, August 2021, and January 2022 (periods just before the Alpha-variant, Delta-variant, and Omicron-variant waves began, respectively). With the objective to demonstrate the forecasting capacity of FIGI-Net, the second column presents the forecast average over the same time time periods. Finally, the last column shows the relative RMSE incurred by FIGI-Net. FIGI-Net successfully predicted the COVID-19 activity shown in the 7-day ahead prediction maps compared to the observation maps. However, some counties had larger errors, as observed from the relative RMSE maps (the last column of Figure 6). During these time periods, counties in Kansas and Louisiana displayed higher errors despite low or mild infection levels. Moreover, the counties in Oklahoma, Iowa, Michigan, and Florida exhibited larger prediction errors preceding the Delta-variant wave. Particularly in Florida, while the epidemic situation was severe, errors increased. Our model demonstrated higher accuracy and lower RRMSE values in the west coast and northeast regions during these time periods (see, for example, the third column of Figure 6, while mid-west and south regions of the U.S. tended to display higher errors as the pandemic progressed (refer to Movie S.1 for the details of geographical distribution prediction and error maps in 1-day, 7-day, and 14-day ahead across all time periods from April 2020 to April 2022).

**Figure 6:**
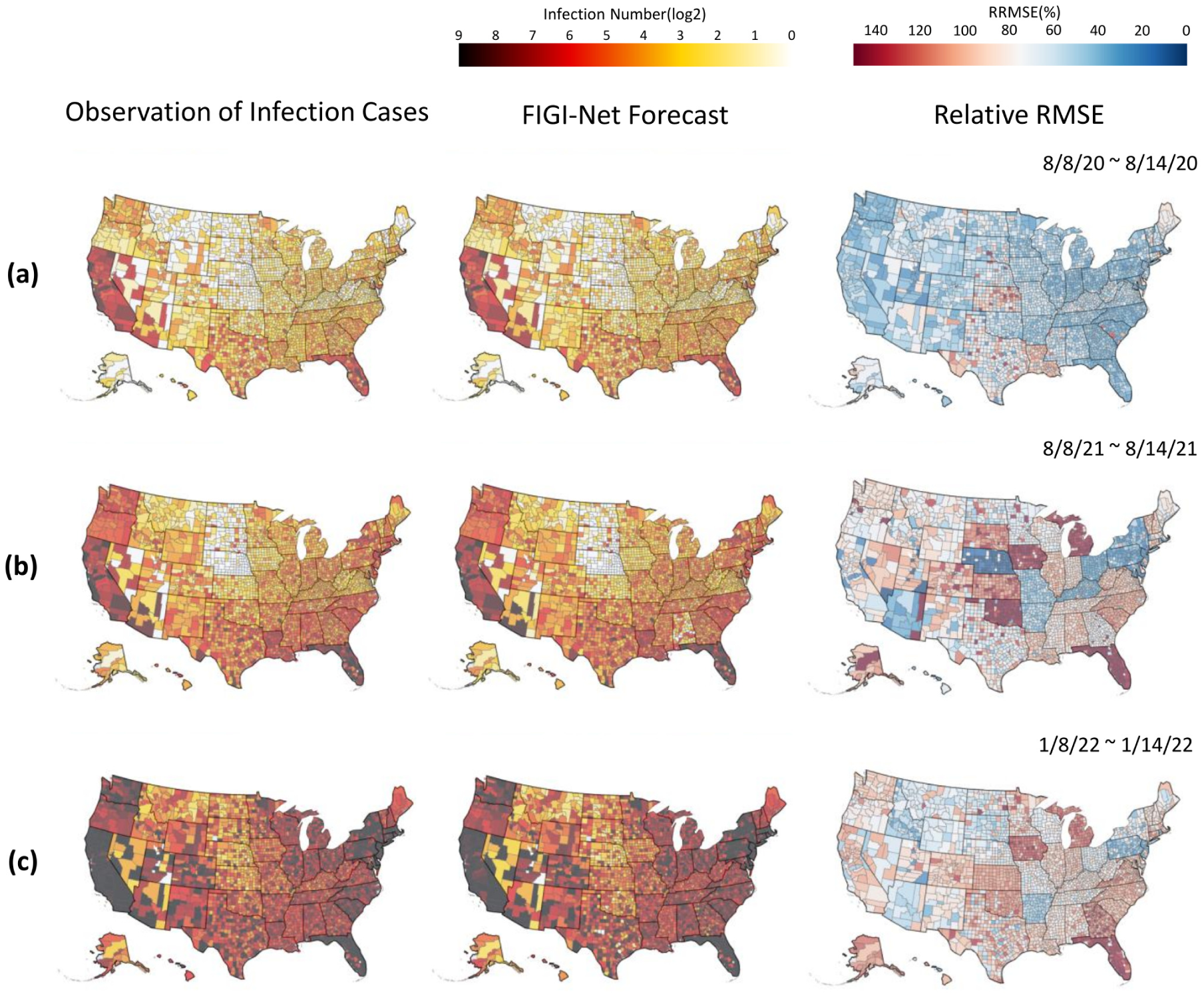
Geographical distribution of infection prediction errors of the proposed model during different time periods. The figure shows the infection prediction distribution of the proposed model during different time periods at county level. The early half of August 2020, August 2021, and January 2022 are represented in (a), (b), and (c), respectively. The first column of (a), (b), and (c) show the observed daily confirmed cases. The second column represent the 7-day (1-week) ahead prediction results, and the corresponding RRMSE score maps are shown in the last column. The proposed model provided the predictions that are similar to the observed daily reports of infection cases and had low RRMSE values in most counties. However, it had higher RRMSE values in the counties where the number of infection cases rapidly increased. Additionally, our model indicated higher RRMSE values in the counties of Kansas and Louisiana at these time periods. The counties in Michigan and Florida represented much higher RRMSE values during time periods (b) and (c) when the status of infections in these two states were severe.

### Comparison between FIGI-Net and the CDC Ensemble Model in COVID-19 Forecasts

To further evaluate our approach, we compared the performance of our proposed FIGI-Net model with the COVIDhub ensemble model [11] (also known as the CDC model). The CDC model employs an ensemble methodology that combines the output of several disease surveillance teams across the United States, generating forecasts for the number of COVID-19 infections at the county, state and national levels. We collected the 1-week and 2-week ahead forecasts of the CDC model at county and state level, and compared them against FIGI-Net’s forecasts. Given the CDC model is an aggregated forecast (i.e. the total number of reported activity over the next 7 and 14 days, rather than a daily forecast over the same periods), we aggregated our daily predictions for the 1 to 7-day and 1 to 14-day horizons to facilitate a fair comparison between both models. A Persistence model of this task was also included as baseline.

Shown in Figure 7.A, the CDC model exhibited significantly higher average RMSE and RRMSE values at the county level compared to our FIGI-Net model. Our model achieved an approximate reduction of 58.5% in averaged RMSE and 53.28% in averaged RRMSE over the 1 and 2-week ahead horizons, respectively (see Table 4). At the state level (Figure 7.B), our proposed model consistently maintained the averaged reduction of 64.55% RMSE value and 64.48% RRMSE value (compared to the CDC ensemble and Persistence models, as shown in Table 5). Notably, the CDC model demonstrated better performance than the Persistence model in terms of lower error predictions for both 1 and 2-week horizons.

**Figure 7:**
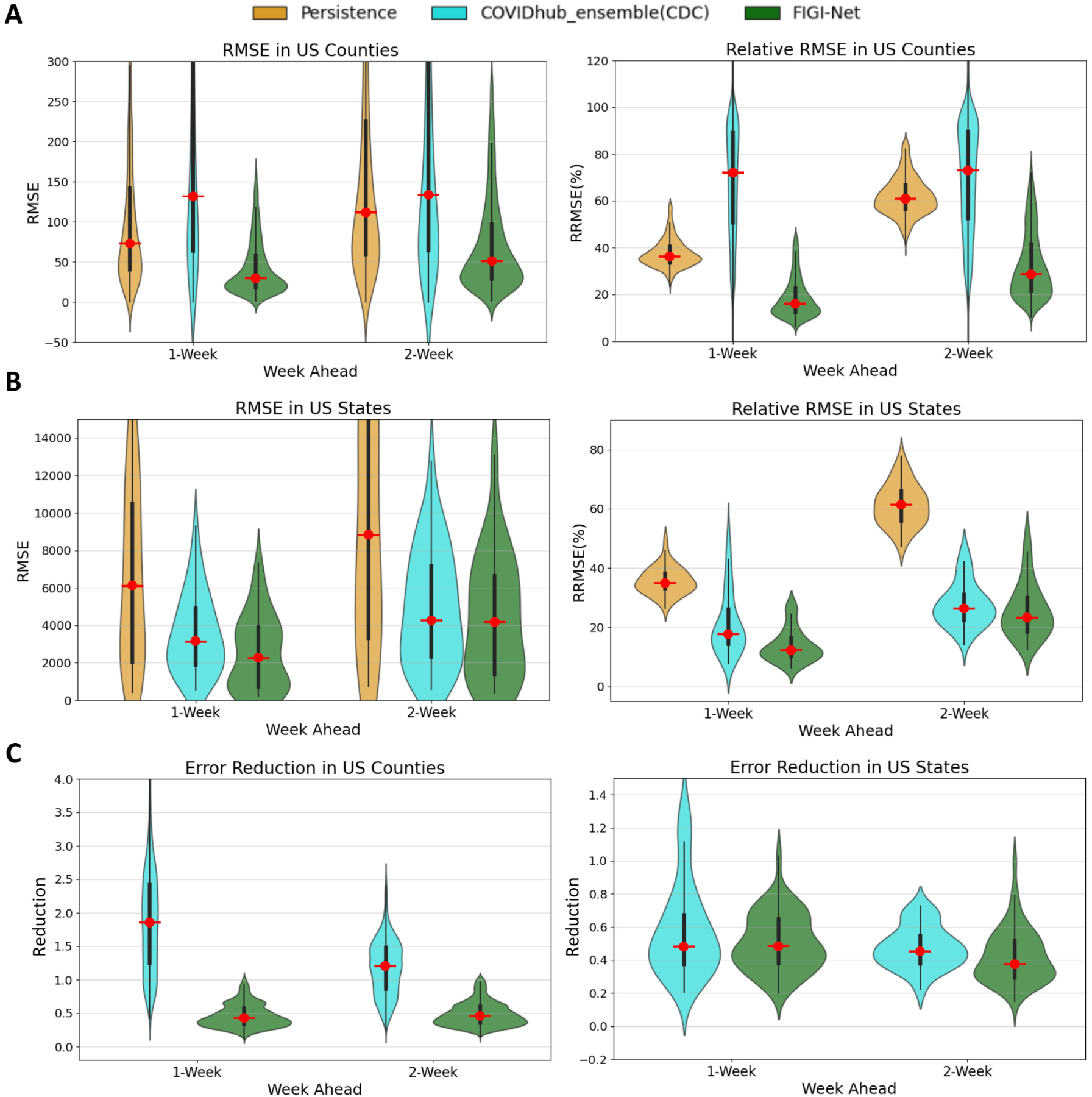
Comparison of weekly COVID-19 infection forecasting performance among Persistence model, CDC model, and our proposed model. (A) The RMSE and relative RMSE values of the three models at the county level. (B) The comparison of prediction errors among these three models at the state level. Our proposed FIGI-Net model outperforms the Persistence model and CDC model in terms of lower prediction RRMSE errors. This indicates its enhanced capability to capture the complex dynamics of COVID-19 infection spread, with approximate 4.76% averaged reduction in errors observed across various prediction horizons. (C) The error reduction comparison between the CDC model and FIGI-Net. At the county level, FIGI-Net outperforms the CDC model, with errors approximately 58.5% lower than those of the Persistence model. At the state level, FIGI-Net continues to provide a 13% lower error reduction compared to the CDC model.

**Table 4:** Comparison of FIGI-Net with CDC and Persistence Models at County Level.

| Model | 1 week |  |  |  | 2 week |  |  |  |
| --- | --- | --- | --- | --- | --- | --- | --- | --- |
|  | RMSE | RRMSE(%) | Error Reduction | P-value | RMSE | RRMSE(%) | Error Reduction | P-value |
| Persistence | 81.24±302.06 | 37.38±14.77 | — | 0 | 125.12±510.7 | 61.67±11.62 | — | 0 |
| CDC_ensemble | 157.49±1285.95 | 76.24±78.71 | 4.63±2.59 | 0 | 157.58±1090.97 | 76.49±66.24 | 2.83±1.98 | 0 |
| FIGI-Net | **34.16±190.24** | **17.13±16.49** | **0.36±0.3** | — | **58.43±282.14** | **29.36±21.21** | **0.47±0.3** | — |

**Table 5:** Comparison of FIGI-Net with CDC and Persistence Models at State Level.

| Model | 1 week |  |  |  | 2 week |  |  |  |
| --- | --- | --- | --- | --- | --- | --- | --- | --- |
|  | RMSE | RRMSE(%) | Error Reduction | P-value | RMSE | RRMSE(%) | Error Reduction | P-value |
| Persistence | 6969.06±11363.25 | 35.65±11.22 | — | 8.65×10<sup>-9</sup> | 11019.9±20085.45 | 62.27±9.51 | — | 2.1×10<sup>-9</sup> |
| CDC_ensemble | 3275.07±4823.08 | 18.93±22.94 | 0.48±0.23 | 0.0021 | 5112.93±8774 | 26.7±19.18 | 0.49±0.16 | 0.0233 |
| FIGI-Net | **3053.46±8315.92** | **12.54±11.29** | **0.34±0.36** | — | **4519.95±10700.57** | **23.57±12.97** | **0.37±0.29** | — |

We also conducted comparative analyses between the COVID-19 forecasts of FIGI-Net with forecasting models officially reported on the CDC website [11] (we selected models, including Microsoft-Deep, JHU CSSE DECOM, Karlen-pypm, CovidAnalytics-DELPHI, and MIT ISOLAT-Mixtures, that provided sufficient infection prediction outcomes for 1-week and 2-week ahead horizons). Following the CDC weekly reporting criteria [1], we aggregated the daily prediction cases into weekly prediction values. Specifically, we focused on three reported infection wave time periods and presented 1-week and 2-week prediction results of our model at the national level alongside those of other models (Figure 8.A). Our findings revealed that most models can accurately predict infection numbers during decreasing trends, but struggle to forecast the correct trend direction and COVID-19 official infection numbers during increasing trends. Interestingly, our FIGI-Net model correctly predicted the increasing direction of the infection trend before the wave began, from November 2021 to March 2023. Figure 8.B illustrates the examples of the predicted infection trends in Massachusetts and New York states by our FIGI-Net model and other forecasting models in 1-week and 2-week horizons. The results showed that the our model’s infection prediction trends are much closer to the reported data across different weeks ahead at the state level, and most of other models predicted accurate infection numbers when the infection trend increases. It is important to note that some models did not provide outcomes at some weekly time points, leading to 0 values in those models, which were subsequently removed in further comparison and analysis.

**Figure 8:**
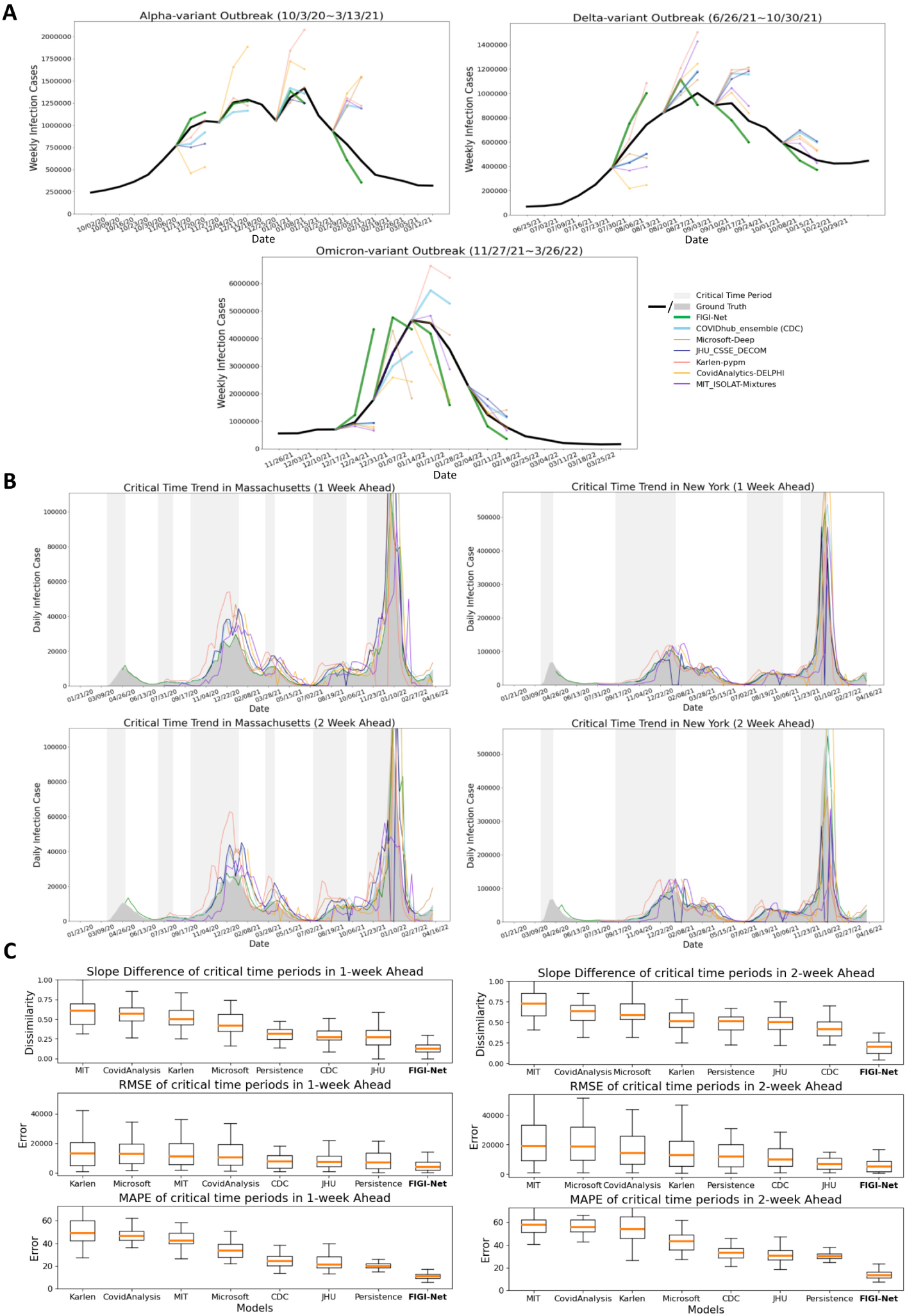
Comparison of FIGI-Net model with other state-of-the-art prediction models in predicting weekly and critical time infections. (A) Comparison of the forecasting results among different forecasting models during three different outbreak time periods at the national level. (B) Examples of COVID-19 infection prediction trends of different state-of-the-art forecasting models at the state level. The critical time period, which indicates a significant increase in COVID-19 infections, is highlighted in light grey color. (C) Performance evaluation of the forecasting methods during the critical time periods of COVID-19 infection in 1-week and 2-week horizons across the states. Slope Similarity, RRMSE, and MAPE were measured to assess the prediction number and trend accuracy of each model. Our proposed FIGI-Net model provided lower prediction errors in both 1-week and 2-week horizons during the critical time and may efficiently forecast the infection number and trend direction before the severe transmission of COVID-19. Here we also ranked them from high to low evaluation or error values according to the median values.

### Comparative Analysis of COVID-19 Forecasting Models During Critical Time Periods

Given that identifying the beginning of a major outbreak is a crucial task in disease forecasting, we assessed the performance of our FIGI-Net model in early COVID-19 prevention and forecasting by measuring its ability to anticipate critical time periods marked by exponential growth of COVID-19 infection cases. We compared our model’s performance with other state-of-the-art COVID-19 forecasting models during this critical time period, using weekly data from July 20^th^, 2020 to April 11^th^, 2022. First, we identified these ”critical” time periods as periods where the trend *λ* of COVID-19 activity (estimated as the coefficient of a linear model *y_t_* = *λy_t−_*_1_) remained above 1, indicating a sustained multiplicative growth for an extended period (Figure S5 to S7 for more details). Examples of such periods for Massachusetts and New York states are shown in Figure 8.B. We compared the ground truth and predicted results of the models using slope similarity, RMSE, and MAPE values (see Section 4.D for a definition to calculate slope similarity). Our FIGI-Net model efficiently and accurately predict infection case numbers and trend direction for 1-week and 2-week horizons during critical time periods (Figure 8.C). Table 6 and 7 presented the forecasting performance details among the prediction models and the statistical significance between our model and the others. Comparing with Persistence model, FIGI-Net model improved the RMSE value by at least 43% reduction in 1-week ahead and at least 57% reduction in RMSE in 2-week ahead forecasts. Additionally, our model achieved at least a 45% MAPE reduction in both week horizons and exhibited around 83% similarity in the slope of infection trend. These results indicate that FIGI-Net model can effectively adapt and refine forecasting trends when the pandemic intensities suddenly.

**Table 6:** Performance of different forecasting models during critical time periods in 1-week ahead.

| Model | Persistence | FIGI-Net | CDC | Karlen | CovidAnalysis | JHU | MIT | Microsoft |
| --- | --- | --- | --- | --- | --- | --- | --- | --- |
| Slope Dissimilarity | 0.316±0.079 | **0.127±0.076** | 0.273±0.108 | 0.5±0.147 | 0.57±0.136 | 0.269±0.134 | 0.606±0.166 | 0.421±0.137 |
| RMSE | 7015.59±15176.43 | **3964.97±6690.04** | 7773.23±14058.41 | 13056.12±14803.5 | 10394.45±21478.21 | 7241.44±9418.22 | 11170.42±22911.14 | 12775.19±26537.04 |
| MAPE | 19.96±3.7 | **11.15±2.97** | 24.48±27.51 | 49.08±76.09 | 46.45±23.76 | 21.07±57.56 | 42.64±29.46 | 33.64±47.7 |
| P-value(v.s. FIGI-Net) | 1.99x10<sup>-9</sup> | - | 4.89x10<sup>-8</sup> | 9.31x10<sup>-10</sup> | 5.15x10<sup>-10</sup> | 1.92x10<sup>-6</sup> | 5.45x10<sup>-13</sup> | 5.15x10<sup>-10</sup> |

**Table 7:** Performance of different forecasting models during critical time periods in 2-week ahead.

| Model | Persistence | FIGI-Net | CDC | Karlen | CovidAnalysis | JHU | MIT | Microsoft |
| --- | --- | --- | --- | --- | --- | --- | --- | --- |
| Slope Dissimilarity | 0.316±0.079 | **0.205±0.093** | 0.419±0.116 | 0.515±0.147 | 0.635±0.138 | 0.501±0.12 | 0.727±0.182 | 0.588±0.172 |
| RMSE | 12141.1±25299.09 | **5126.55±9211.25** | 10087.22±22735.07 | 13032.49±17830.7 | 14211.7±30659.77 | 6969.87±10211.86 | 19078.33±40946.07 | 18664.36±38065.72 |
| MAPE | 30.05±3.98 | **13.28±3.9** | 33.38±19.77 | 53.98±78.8 | 55.88±21.28 | 30.63±51.35 | 57.81±24.24 | 43.47±44.71 |
| P-value(v.s. FIGI-Net) | 3.73x10<sup>-9</sup> | - | 7.74x10<sup>-9</sup> | 3.94x10<sup>-9</sup> | 1.57x10<sup>-9</sup> | 0.001 | 1.32x10<sup>-11</sup> | 1.25x10<sup>-9</sup> |

## Discussion

In this work, we have introduced FIGI-Net, a deep learning-based model that utilizes fine-grained county level infection time-series data for short-term forecasting up to two weeks. We evaluated the forecasting ability of FIGI-Net against state-of-the-art methodologies, including autoreggressive models, recurrent network architectures such as GRU and LSTMs, and more advanced deep-learning architectures such as TC-LSTM and bidirectional LSTMs. Our strictly out-of-sample analysis, from October 18^th^, 2020 to April 15^th^, 2022, shows that FIGI-Net represents an improvement over existing state-of-the-art models, successfully predicting COVID-19 dynamics at the county, state, and national levels, across multiple time horizons, reaching error reductions of up to 90%, 89.3% and 86.48% in RRMSE, accordingly.

At the county level, FIGI-Net successfully predicted COVID-19 activity, scoring error reductions of up to 90% in comparison to the baseline model, Persistence. FIGI-Net consistently placed as a top 1 performer across the multiple time horizons based on error metrics (RMSE and RRMSE), as presented in Table 1. At the state level, we compared FIGI-Net predictions against state-of-the-art models for the 1-day, 7-day and 14-day horizon. Our results showed that FIGI-Net achieved error reductions of 89.3%, 53.76%, and 41.1539.42% in RMSE accordingly, when compared to the Persistence estimate (see Table 2). Finally, at the national level, our model successfully presented an error reduction of up to 86.48% over the Persistence estimates. We attribute the success of FIGI-Net across multiple geographical resolutions to the pre-trained model component in our framework, which captures meaningful patterns across global infection dynamics. This clustering-based approach utilizes global spatio-temporal features learnt a priori, enabling subsequently fine-tuned sub-models to mitigate the influence of noise or irrelevant information and increase its own predictive power. This innovative framework better captures the rapidly changing dynamics (see, for example, Figure 3), ensuring accurate forecasts up to 2 weeks into the future.

Furthermore, we conducted training on several models exclusively utilizing State-level data and compared the results with those derived from our model, as shown in Figure S1. According to this comparative analysis, the deep learning-based model requires a sufficient amount of effective data size to help reinforce its response to sudden changes in forecasting. Leveraging County-level data provides an ample amount of training information to generate predictive outcomes at a small scale, which can then be aggregated to yield enhanced precision in forecasting at a coarser scale.

The analysis of FIGI-Net predictive power against the CDC’s official ensemble predictions showed notable improvements seen as substantial reduction in error rates produced by FIGI-Net. At the county level, our model demonstrated more than 50% error reduction, while at the state level, the reduction was at least 63% (see Table 4 and 5). This success is indicative of our model’s adaptation capacity, particularly at the granular county level — a domain where the CDC’s ensemble predictions exhibited comparatively poorer performance. This suggests the potential of our model not only in refining county-level forecasts, but also in addressing the nuances that contribute to more accurate forecasting, highlighting its utility in augmenting current predictive methodologies.

The forecasts from FIGI-Net presented in this work were created using a training moving window consisting of 75 days of data in length. Our choice of training window is based on an experimental analysis on the predictive power of FIGI-Net as a function of the training window size. Figure 2 illustrates that longer length of training data positively influenced our model’s performance, as assessed and identified through the initial six-month dataset for robust evaluation. Particularly, there is a small change in 2-week horizon’s performance when the training data length exceeded 75 days, whereas a longer training data length shows enhanced performance for a 1 week horizon. Based on our 1 and 2 week horizon’s performances (presented in Figure 7), we determined the optimal training data length to be 75 days. This time period is short enough to capture changes in disease transmission in a responsive way.

Upon analyzing the geographical distribution maps presented in Figure 6, which illustrate our county-level predictions and errors across the U.S., it is evident that variations in reporting values might arise from changes in epidemic prevention policies in different regions. These differences in data format can significantly complicate the accurate prediction of COVID-19 infections and lead to a substantial increase in errors. For example, during the Delta-variant wave, as shown in Figure 6(b), the RMSE for counties in Kansas exceeded that of neighboring states, even though the pandemic risk in those areas was relatively mild. This pattern is also noticeable in Iowa during the Omicron-variant wave, as depicted in Figure 6(c). We attribute these observations to two factors: (1) the relationship between low daily reported infection cases and higher predicted outcomes, leading to larger errors. For instance, if a county reports 2 infection cases while the prediction is 4 cases, this discrepancy results in a larger RMSE. (2) State governments change their recording policies from daily to weekly at certain periods, introducing inconsistencies and irregularities in the data format that could impact model predictions. This policy change points out the importance of consistent instructions and practices for specific epidemiological diseases, ensuring effective management of public healthcare information and promoting accurate disease analysis, prediction, and prevention. Further studies are necessary to explore and address these issues in order to enhance the accuracy and reliability of infection predictions.

As depicted in Figure 8.A and B, most assessed forecasting models struggled to predict the direction of future infection trends accurately during multiple time periods, perhaps due to the highly variable transmission rates of the multiple COVID-19 variants. Importantly, FIGI-Net predicted appropriately the trend direction during the initial days of each of the three outbreaks that were studied. This capability is attributed to our model’s daily prediction of infection case numbers, which has allowed for the early detection and response to sudden changes. Furthermore, utilizing county-level data with clustering allows the identification of early regional variations and swift adjustment of the forecasting trend by the proposed sub-models. These features enable our model to efficiently adapt to dynamically various changes in infection numbers and trends during COVID-19 outbreaks. Moreover, our model exhibited a higher slope similarity score (See Table 6 and 7), lower RMSE and MAPE scores than others. These results indicate that the proposed model excels at predicting the direction of forecasting trend, facilitating early implementation of COVID-19 transmission prevention. Our study underscores the robustness and effectiveness of time-series deep learning-based methods in handling dynamic and sudden changes in infection numbers during short-term time periods.

Based on the experiments, our proposed model has certain limitations. Firstly, our deep learning-based model requires a longer training time, compared to linear models, due to the complexity and computational demands. Although our model can automatically obtain optimal hyperparameters, this leads to extended convergence times. Additionally, each cluster has its trained model to enhance forecasting outcomes, but this increases computational time for predicting a single time period. Another limitation is the requirement for an adequate amount of training data. Deep learning models need large volumes of diverse and representative data to learn underlying patterns effectively and make accurate predictions. Limited data can compromise the model’s performance. Therefore, ensuring a substantial amount of high-quality training data is essential for our model’s effectiveness. This could be achieved by increasing the granularity of infection case data, such as building town-level datasets. Addressing these limitations is crucial for the model’s real-world application. Future research ought to overcome some of these challenges.

In conclusion, the FIGI-Net model represents an improvement in the field of COVID-19 infection forecasting and may serve as a template for future pandemic events. By employing temporal clustering and a stacked structure of biLSTM, our model achieves accurate and efficient COVID-19 infection forecasts from fine-grained county level datasets. Accurate and early predictions of COVID-19 outbreaks at the county, state, and federal levels is of paramount importance for effective public health management. Our model’s ability to provide early warning of potential outbreaks allows prompt and targeted public health interventions. The potential applications of our model in public health management and epidemiological disease prevention are substantial and could profoundly impact mitigating the effects of future infectious disease outbreaks.

## Data and Methods

### Data Collection and Cleaning

The data utilized in this study includes the daily COVID-19 cumulative infectious and death cases of U.S. counties, obtained from the Johns Hopkins Center for Systems Science and Engineering (CSSE) Coronavirus Resource Center between January 21^st^, 2020 and April 16^th^, 2022[15]. It is important to note that each county or state government may have different policies for pandemic recording and reporting, which can make the CSSE data difficult to evaluate and analyze. Additionally, cumulative data may not efficiently differentiate regional variation. To address these issues, we utilized a 7-day average method to denoise the daily official COVID-19 cases [35]. We average the case number of the current day and the preceding six days to obtain the denoised value. Because CSSE data occasionally had abnormal or missing-data observations, we selected valid data, including only counties within the continental United States and verified confirmed case data, of U.S. counties, including 3143 counties between February 2020 to March 2022, as the ground truth for further evaluation. Due to the rapid changes in the COVID-19 situation, we partitioned the dataset into 48 time intervals, each spanning approximately 15 days in length, to train our models separately.

### Temporal Clustering

Based on the evidence that neighboring COVID-19 dynamics may highly influence local dynamics (see Noor *et al* [43]), our methodology incorporates a two-step spatio-temporal clustering procedure that aims to identify both global and local similarities between the COVID-19 activity within each county in the U.S.. The outcome of this analysis guides our framework to train sub-models that are fit only on the most relevant dynamics for each county. The procedure consists in the following steps: 1) Creating a set of feature vectors representing the similarity between each county, 2) Compressing the representation of this features via a dimensionality-reduction procedure (in this case, we apply UMAP), and 3) A 2-level clustering procedure over the resulting lower dimensional version of combined correlation feature vectors.

#### Creating the feature vectors

For each time period, we computed the autocorrelation [53] of COVID-19 daily confirmed cases and fatality rates, and obtained the cross-correlation between these two data. These two correlation features were concatenated per county, resulting in feature vectors representing the combined correlation information for each county.

#### Dimensionality reduction

Given our feature vectors are of dimension 301 given the number of counties within the us, a lighter representation of this feature vectors is necessary. We transformed our feature vectors using the UMAP method [39]. UMAP is dimensionality reduction technique to effectively prevent and handle global and local nonlinear structure in a lower-dimensional space. We selected UMAP as our dimensionality-reduction step given its ability to make an optimal choice for preserving both local and global relationships during the reduction process. The resulting outcome in this step is a feature vector of dimension 2.

#### 2-level clustering

To identify temporal clusters, we used the unsupervised DBScan method [18] to extract initial clusters, followed by selecting the largest cluster to obtain the subclusters via spectral clustering [41]. Figure 9 shows a representative example illustrating the process of temporal clustering. The initial clustering identifies the global differences in county trends, and the subclustering distinguishes local differences from similar reporting trends. By seeking similar temporal features, we identified the clusters that include counties from different geographical locations, enabling the provision of relevant local information. Subsequently, the list of cluster labels was rearranged in descending order of infection risk based on the average infection count over last 7 days within each cluster. According to the results of our experiments, we determined the optimal number of clusters to be 8 for facilitating further submodel training. When the number of clusters was too small, the submodel may learn irrelevant infection features that could adversely impact local predictions. Conversely, if we identified too many clusters, the size of training data becomes smaller and may not provide enough training data for submodel training. We found that 8 clusters achieve a good balance between providing relevant local information for accurate predictions and ensuring an adequate amount of training data.

**Figure 9:**
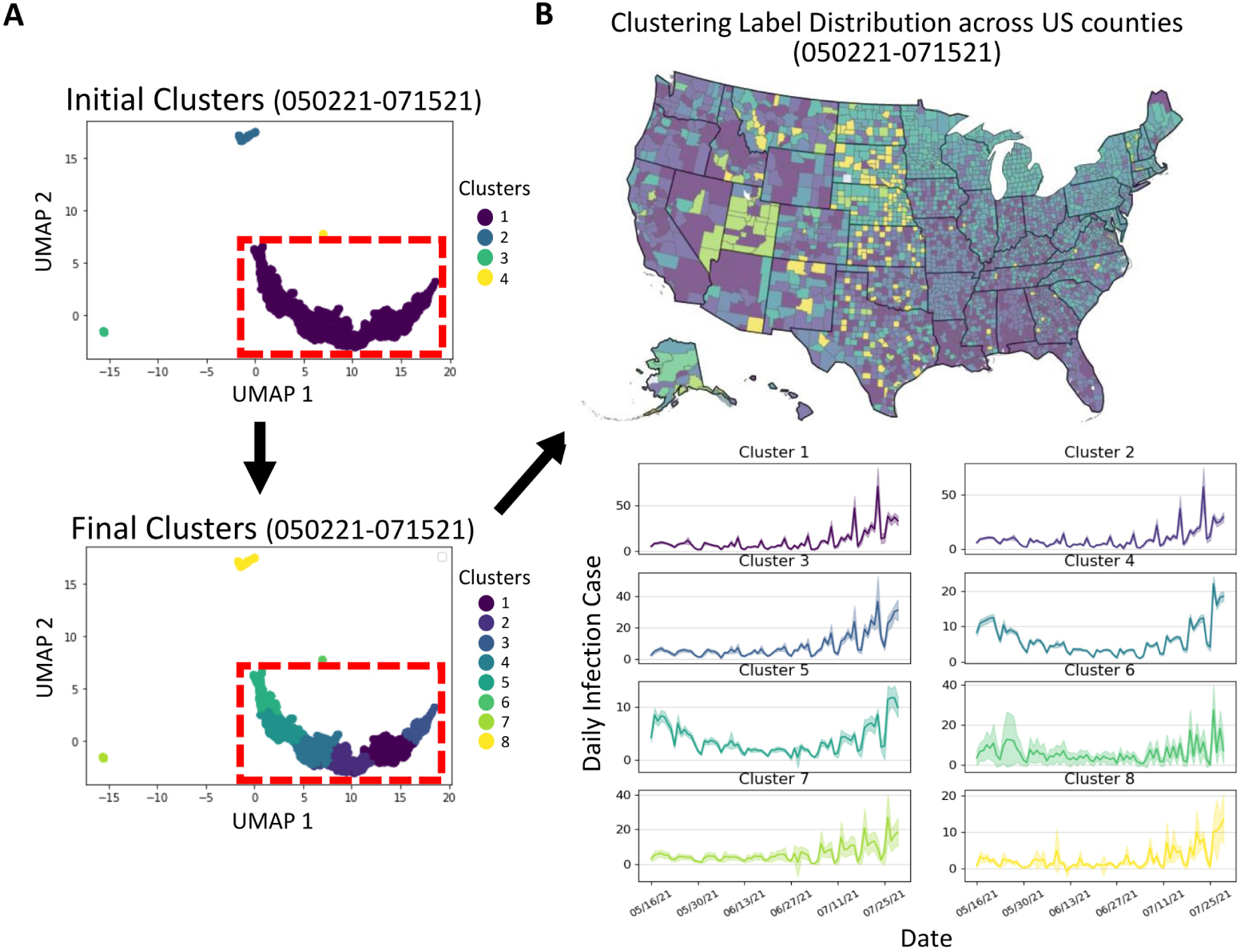
An example of temporal clustering in a specific time period. (A) The UMAP method is utilized to map the correlated temporal features, which are then grouped to form initial clusters. The largest cluster (highlighted by the red-dashed rectangle) is further subdivided to obtain subclusters, and a total of 8 clusters are reordered based on the average infection number of the last 7 days within each cluster. (B) The geographical distribution across US counties exhibits how temporal clustering captures the relationship between time-series and spatial information. The infection curves of the training data (with a duration of 75 days) for the clusters, along with a 95% confidence interval, illustrate that this approach provides a collection of highly related yet distinct subclusters to help the sub-model efficiently learn and make accurate predictions.

### Training sub-models through transfer learning from a global model

Due to the dynamic and rapid changes in the trend of COVID-19, the use of time series forecasting models has become essential. The LSTM model, developed from the recurrent neural network, is particularly useful in handling time series forecasting problems [24]. In order to address the short-term infection variability, we implemented a bidirectional stacked LSTM (biLSTM) model for predicting infection case trends, and the architecture of the proposed model is shown in Figure 1(b). It leverages the bidirectional method to learn the variability of the future sequence trends over time and strengthens the ability to handle unexpected sudden changes. Furthermore, the stacked structure helps our model recognize the similarity and relevance of the entire historical trend of each time period among counties.

To achieve accurate and efficient forecasts in each time period, we introduced transfer learning to deal with the rapid variability and transmission of COVID-19. We collected 75-day length raw data of all counties before the time period for forecasting and used a 60-day length sliding window, consisting of 45-day length for training inputs and 15-day length for predicted labels, to generate the training dataset of all counties and the counties in each temporal cluster, respectively. We used the training dataset to train the global model to learn universal infection features to address possible trend changes. Then we transferred the parameters of this pre-trained global model to train 8 sub-models for each temporal cluster, and each sub-model was fine-tuned by using the county data of each temporal cluster. During each model training, the training data was randomly split into training (80%) and validation sets (20%). The temporal clustering provides highly relevant historical information, enabling the training of local submodels by optimizing the parameters of the global pre-trained model. This approach ensures accurate forecasting of county-level infection numbers. We used the Adam optimization algorithm, with a learning rate of 10*^−^*^3^ and 100 epochs during the training process.

### Model evaluation

Daily forecasting error is assessed using the RMSE and RRMSE, defined as:

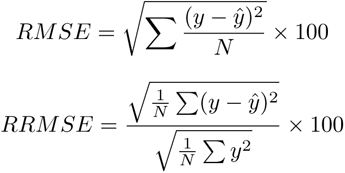

where *y*, *y*^, and *N* represent the observation, the predicted values, and the number of U.S. counties, respectively. Also, to assess the consistency and generalization of the model over time, we use MAPE and error reduction, defined as:

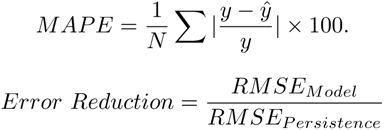

We also evaluated the similarity of slope score, which indicate the difference of trend directions between observations and forecasting models, to measure the accuracy of predicted directions in each time point. The range of slope score is from 0 to 1 and the function is defined as follows,

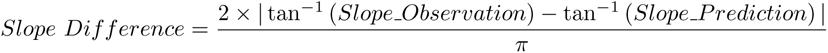

A lower score represents a predicted trend direction that aligns with the ground truth at a specific time point. We employed the linear regression method [19] to calculate the slope of each point by using short time interval, which includes the two points before and after the specific point. These measurement methods were used to assess the accuracy of the forecasting model in reflecting rapid trend changes.

For statistical evaluation, the two-sided Wilcoxon rank sum test, known as the Mann-Whitney U test, was utilized to test statistical significance among the model performances [42]. This testing approach is frequently employed for handling performance metrics without assuming a normal distribution of the data and without specifying the direction of the difference. Additionally, to quantify the variability of forecasting, the function of error range for confidence interval was used and is shown as:

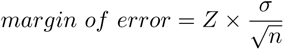

where *σ* and *n* represent standard deviation of samples and the size of samples, respectively. *Z* is set to 1.96 for a confidence level of 95% [42].

## Data Availability Statement

The data used in this study are publicly available and consist of daily COVID-19 cumulative infectious and death cases reported for U.S. counties. The dataset was obtained from the Johns Hopkins Center for Systems Science and Engineering (CSSE) Coronavirus Resource Center, spanning from January 21^st^, 2020, to April 16^th^, 2022 [15]. The dataset can be directly accessed from the Johns Hopkins CSSE Coronavirus Resource Center website (https://github.com/CSSEGISandData/COVID-19). Researchers interested in utilizing the data for further analysis can refer to the original source for detailed documentation on data collection methods and definitions. For additional information or inquiries about the dataset, please visit the website or contact the Johns Hopkins CSSE Coronavirus Resource Center.

## Code Availability Statement

All code used to implement the algorithms and conduct the experiments in this study is available online at the following repository: https://github.com/kleelab-bch/FIGI-Net. Users can access the code, along with the instructions for replicating the experiments and obtaining the forecasting results. All code is shared under the MIT License and can be freely accessed and reused for academic and non-commercial purposes.

## Acknowledgement

K.L. and M.S. were supported by the National Institutes of Health (NIH) under award number R35GM133725. M.S. was also partially supported by the NIH under award number R01GM130668.

M.S. has been funded (in part) by contracts 200-2016-91779 and cooperative agreement CDC-RFAFT-23-0069 with the Centers for Disease Control and Prevention (CDC). The findings, conclusions, and views expressed are those of the author(s) and do not necessarily represent the official position of the CDC.

## Potential Conflicts of interest

M.S. has received institutional research funds from the Johnson and Johnson foundation, Janssen global public health, and Pfizer.

## Notes

### Competing Interest Statement

Mauricio Santillana has received institutional research funds from the Johnson and Johnson foundation, Janssen global public health, and Pfizer. Other authors declare no competing financial or non-financial interests.

### Summary of Updates

The sections of Acknowledgement and Conflict of Interests have been updated. Fig.7 in the previous version was moved to Fig.2 in the revised version.

