## Supplementary Information for "Fine-Grained Forecasting of COVID-19 Trends at the County Level in the United States"

**This PDF File includes:**

- Tables S1 to S4
- Figures S1 to S7

Table S1: Comparison of Forecasting Models with different training format at State Level

| Models | 1st day | | 7th day | | 14th day | |
|---|---|---|---|---|---|---|
| | RMSE | RRMSE(%) | RMSE | RRMSE(%) | RMSE | RRMSE(%) |
| Autoregressive* | 2919.32±20428.46 | 78.63±686.99 | 3600.42±1.31x$10^{13}$ | 101.39±5.41x$10^{11}$ | 15141.28±5.32x$10^{23}$ | 591.01±2.45x$10^{22}$ |
| GRU* | 2614.1±5706.59 | 75.69±38.71 | 2681.13±5703.73 | 76.08±42.74 | 2905.47±5716.06 | 82.43±34.63 |
| LSTM* | 2769.32±5701.13 | 71.69±19.52 | 2764.88±5721.4 | 72.11±18.9 | 2940.21±5707.36 | 79.55±16.32 |
| biLSTM* | 2340.29±5475.5 | 68.51±15.85 | 2521.58±5473.84 | 70.93±16.95 | 2743.3±5483.12 | 77.91±19.24 |
| biLSTM | 371±1047.57 | 12.44±15.04 | 1189.43±1806.38 | 28.9±14.01 | 1940.18±3216.91 | 49.46±17.18 |
| FIGI-Net | **290.31±908.81** | **9.17±13.59** | **1149.66±1850.66** | **26.94±14.48** | **1935.36±3458.21** | **49.12±16.47** |

*: directly use State data for model training

Table S2: Comparison of FIGI-Net against the baseline models in 1 week horizons at the county level.

| Models | RMSE | RRMSE |
|---|---|---|
| Autoregressive | 212.18±2.44x$10^{10}$ | 83.32±4.16x$10^{9}$ |
| Persistence | 81.24±302.06 | 37.38±14.77 |
| GRU | 48.4±254.14 | 26.32±88.23 |
| LSTM | 52.09±240.49 | 26.69±61.04 |
| TC-LSTM | 57.32±281.55 | 28.39±19.87 |
| TC-biLSTM | 40.4±222.56 | 20.27±16.62 |
| biLSTM | 36.63±180.55 | 18.57±57.39 |
| FIGI-Net | **34.16±190.24** | **17.13±16.49** |
| CDC | 157.49±1285.95 | 76.24±78.71 |

Table S3: Comparison of FIGI-Net and baseline models for the 2 week horizon task at the county level

| Models | RMSE | RRMSE |
|---|---|---|
| Autoregressive | 99.79±6.78x$10^{5}$ | 45.25±1.17x$10^{5}$ |
| Persistence | 125.12±510.7 | 61.67±11.62 |
| GRU | 63.89±305.64 | 33.3±116.39 |
| LSTM | 72.64±317.59 | 32.85±48.7 |
| TC-LSTM | 71.7±334.39 | 36.8±25.69 |
| TC-biLSTM | 69.59±311.8 | 34.03±22.64 |
| biLSTM | 59.25±283.61 | 29.88±31.89 |
| FIGI-Net | **58.43±282.14** | **29.36±21.21** |
| CDC | 157.58±1090.97 | 76.49±66.24 |

Table S4: Comparison of training duration identification for the proposed model across 1-week and 2-week forecasting horizons

| Training Length | 1 week | | 2 week | |
|---|---|---|---|---|
| | RMSE | $R^2$ | RMSE | $R^2$ |
| 30 days | 24.45±14.31 | 0.864 | 29.79±11.45 | 0.861 |
| 45 days | 26.28±14.49 | 0.944 | 26.42±13.33 | 0.912 |
| 60 days | 19.04±13.28 | 0.968 | 24.97±14.59 | 0.933 |
| 75 days (Optimal) | 14.39±11.23 | 0.977 | 22.71±12.46 | 0.937 |
| 90 days | 12.74±9.66 | 0.978 | 20.26±12.66 | 0.943 |

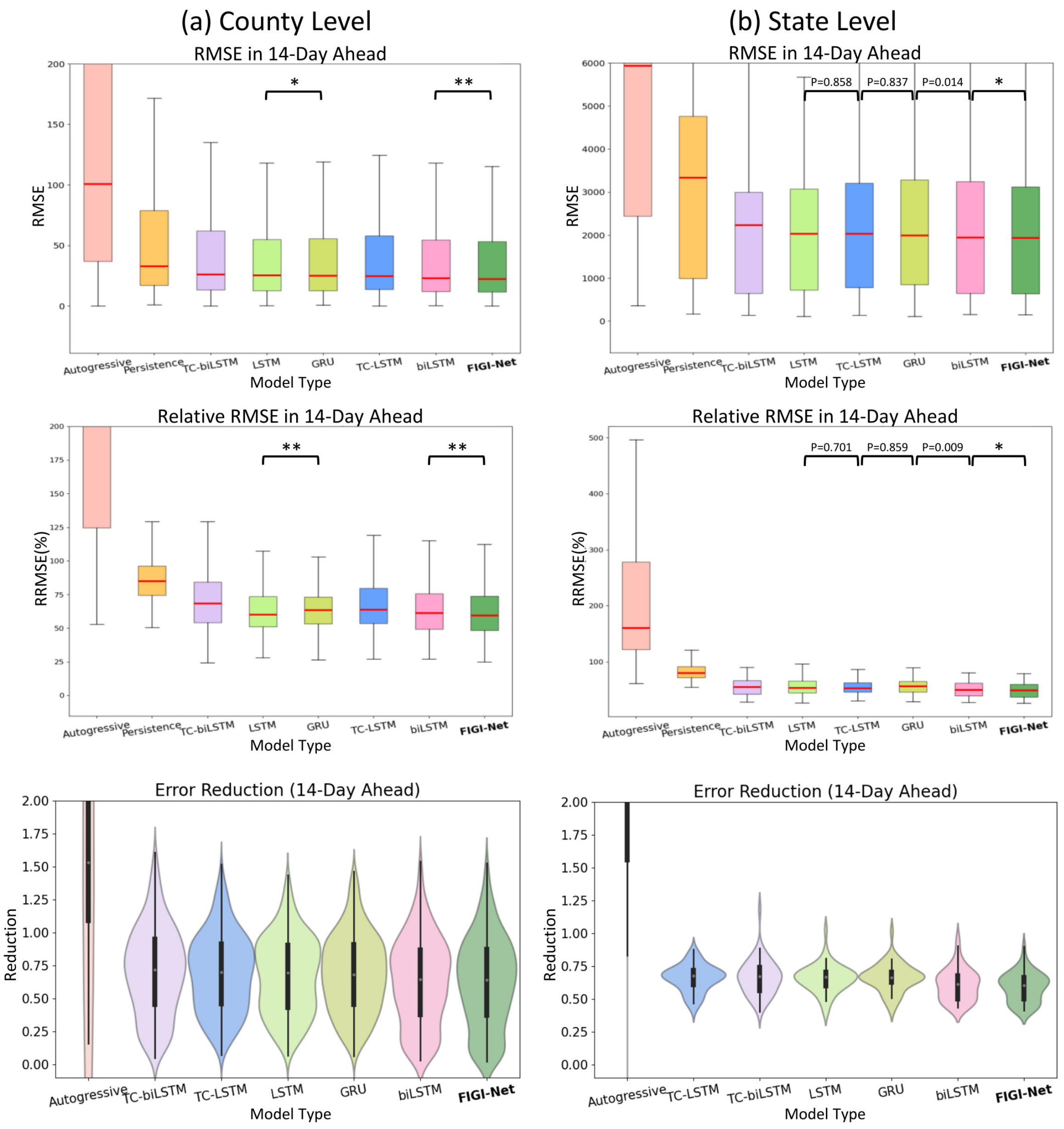


Figure S1: Comparison of Root Mean Squared Error (RMSE), relative RMSE, and error reduction across various models is conducted for a 14-day forecast horizon, analyzed at both (a) county and (b) state levels. The performance assessment reveals that the deep learning-based model exhibits comparable forecasting capability within the 14-day horizon. Furthermore, the FIGI-Net model demonstrates a significant reduction in forecasting errors, approximately 35% at the county level and 50% at the state level, when compared to the persistence model.

: p-value $<0.005$

*: p-value$<0.001$

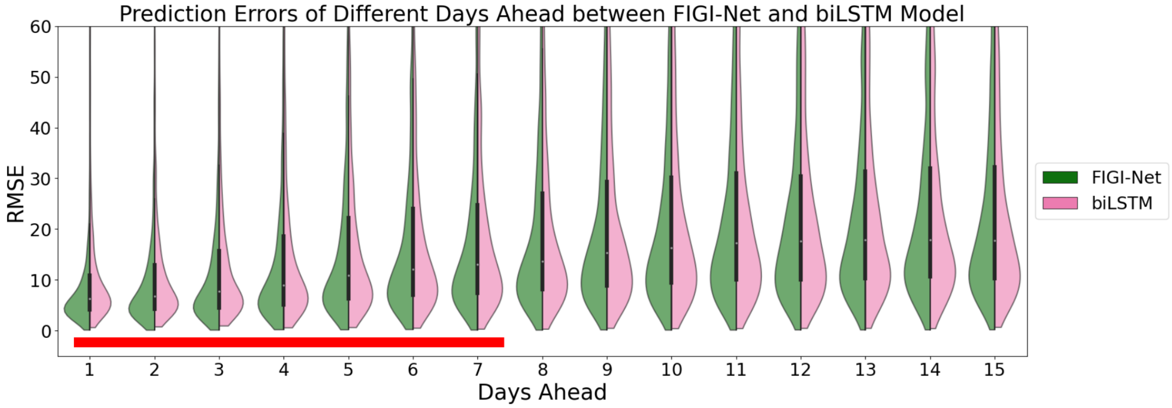


Figure S2: Comparison of RMSE between FIGI-Net and biLSTM across various forecast horizons. Our results indicate that our model yields lower prediction errors for infection numbers within the first 7 days of prediction compared to biLSTM (as indicated by the red line).

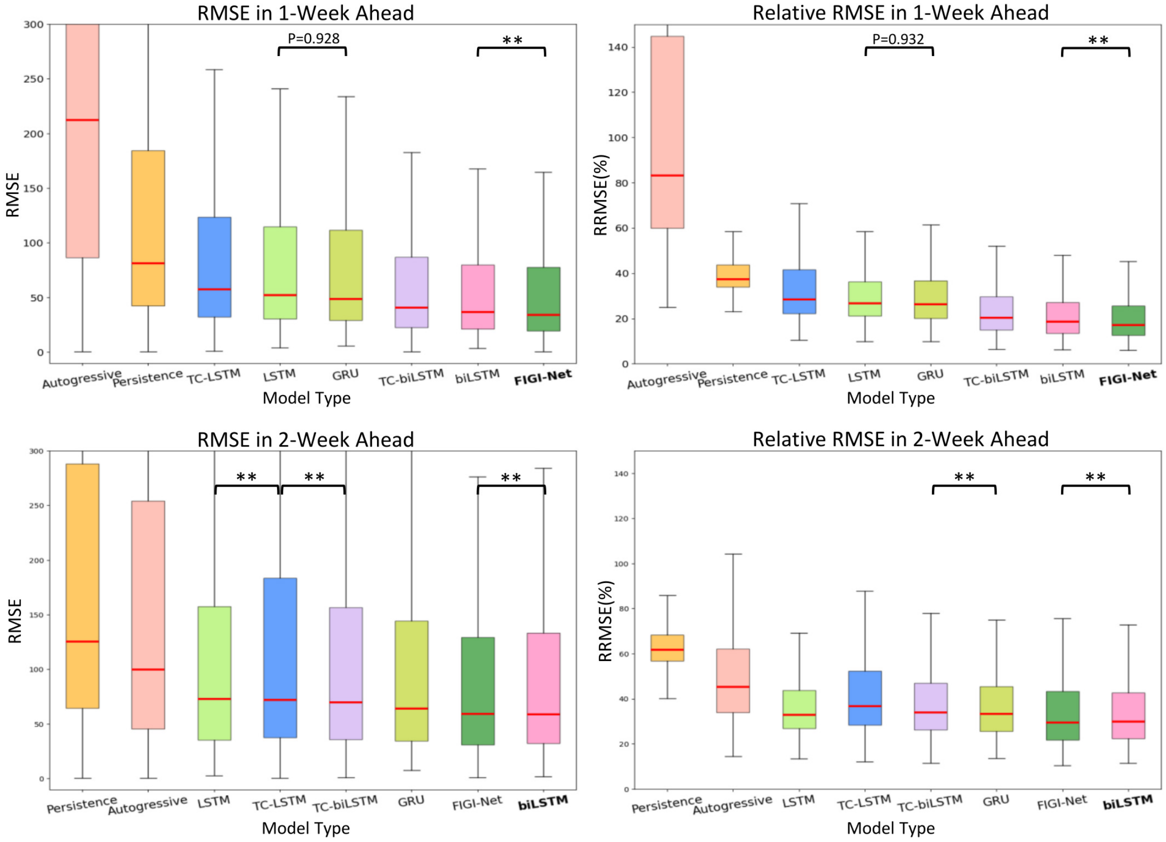


Figure S3: Comparison of various models at the county level across 1 and 2 week ahead horizons. The models are sorted in descending order based on their RMSE values. Notably, our FIGI-Net model demonstrates accurate prediction performance and places among the top 2 with lowest RMSE.
*: p-value<0.001

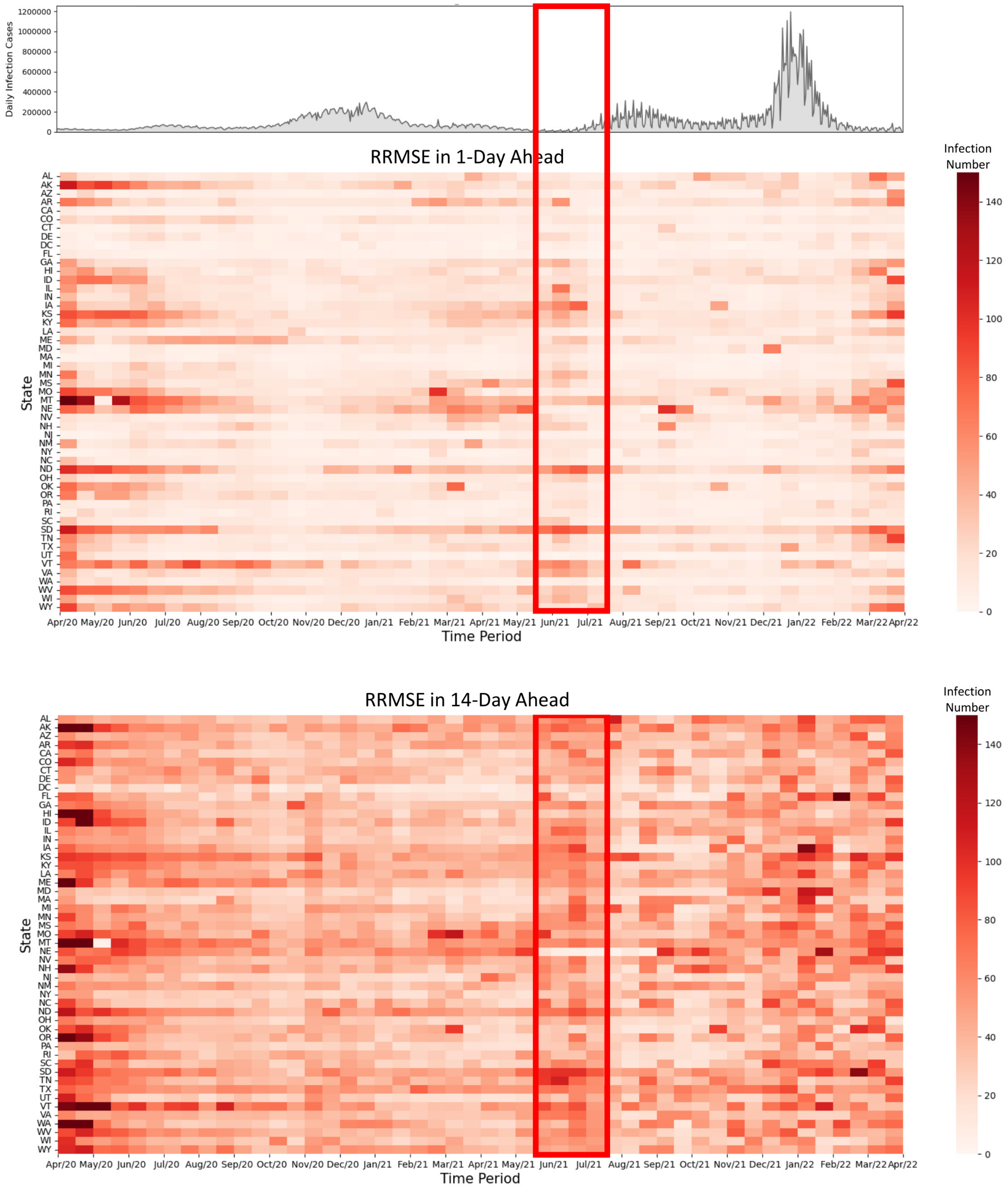


Figure S4: Evolution of prediction error for FIGI-Net at state level. Each column represents the average relative RMSE (each average is computed using a non-overlapping 15-day period between April 2020 and April 2022) across US states for the 1-day and 14-day horizons, compared to the national reported infection trend. We can clearly observe that Missouri, Montana, and Nebraska have large RRMSE values during March 2021 to May 2021 in 1-day ahead prediction. In addition, we can observe that the RRMSE errors increase before the early stage of the next outbreaks (red rectangle).

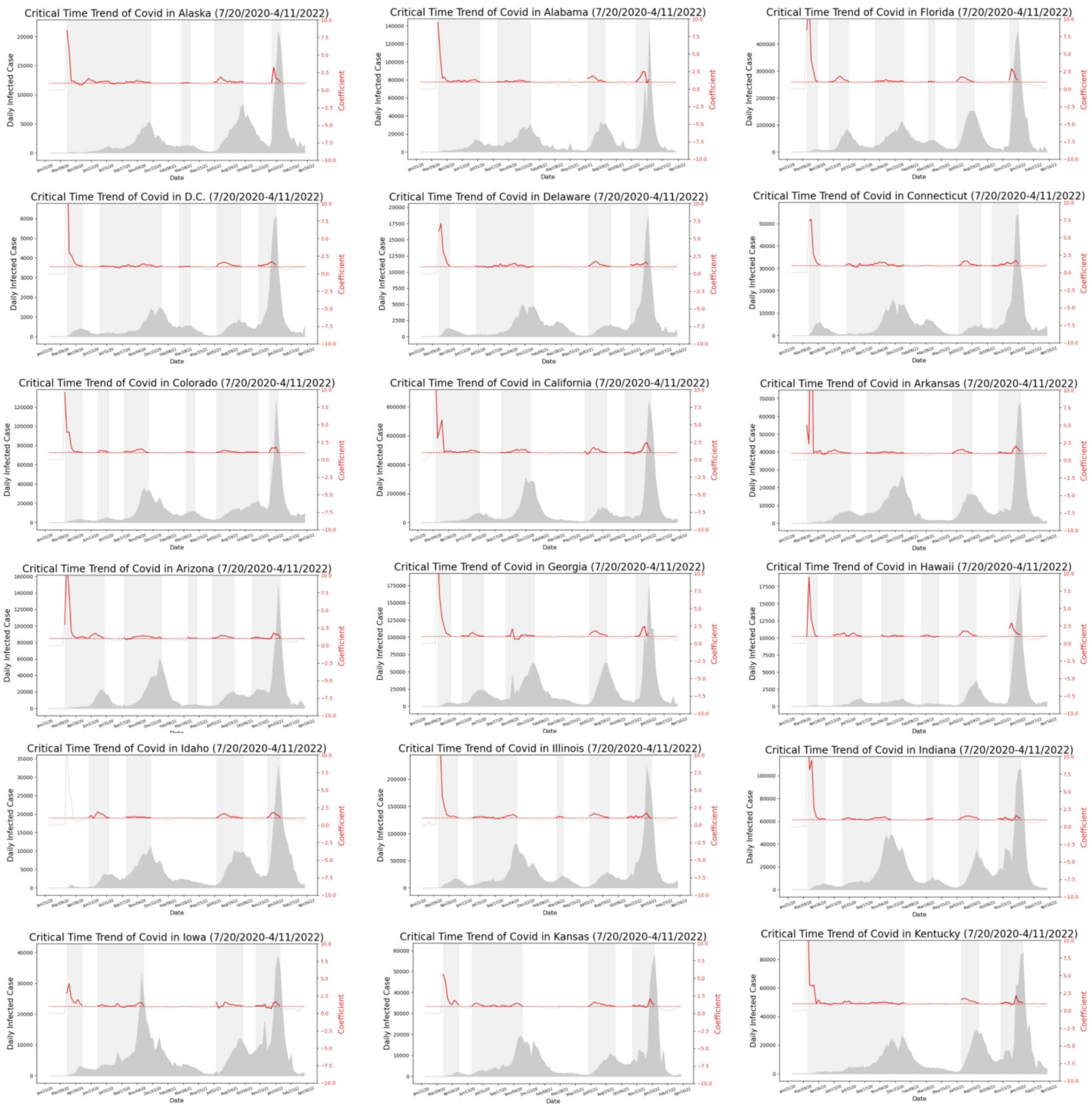


Figure S5: COVID-19 Critical Time Periods for each state in US (Part I). The critical time periods were created by independently quantifying the trend of COVID-19 confirmed cases across each state, and identifying the time periods when the value of the trend exceeded the threshold of 1.0 (marked with a red-dash line in every figure).

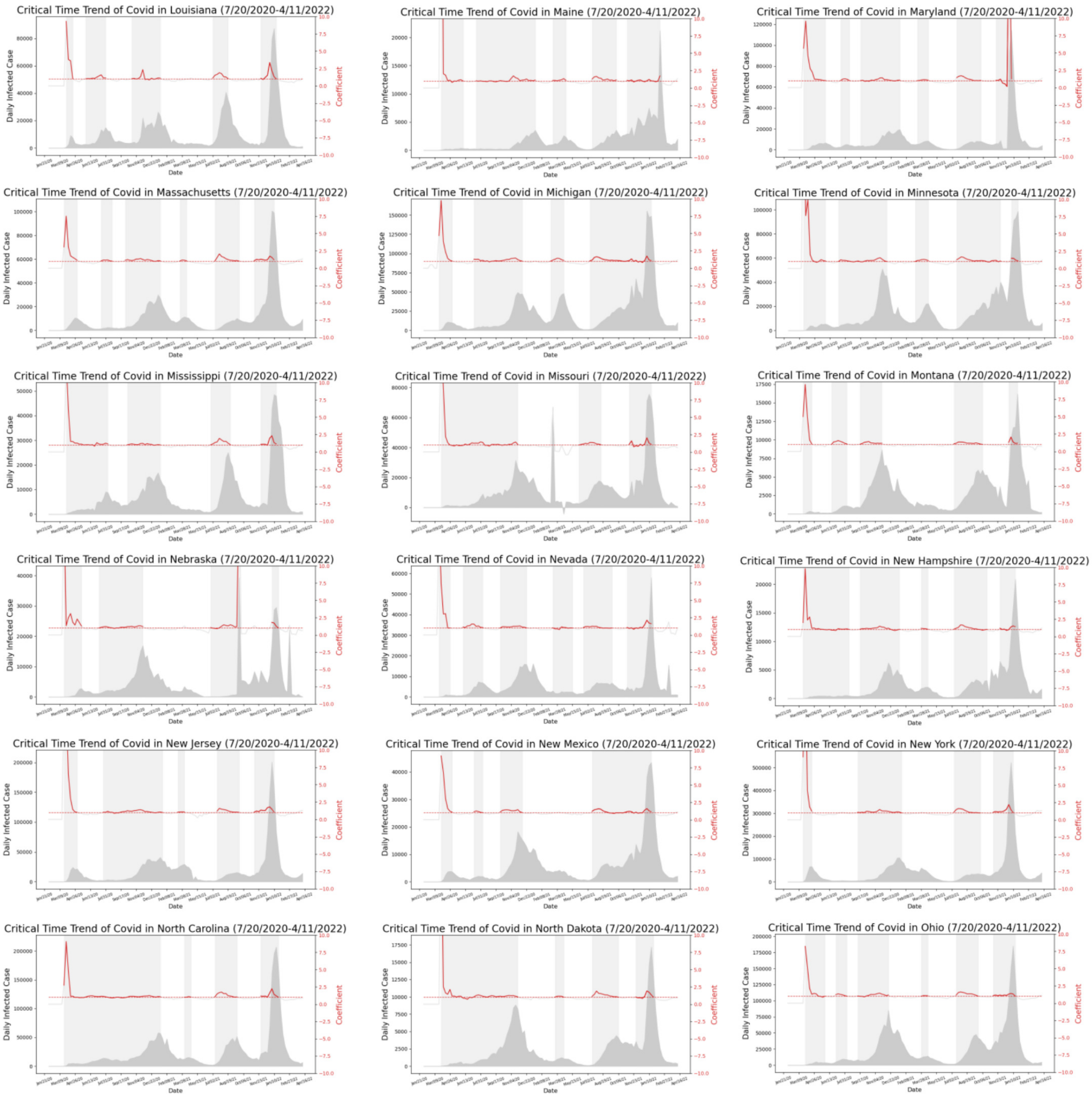


Figure S6: COVID-19 Critical Time Periods for each state in US (Part II).

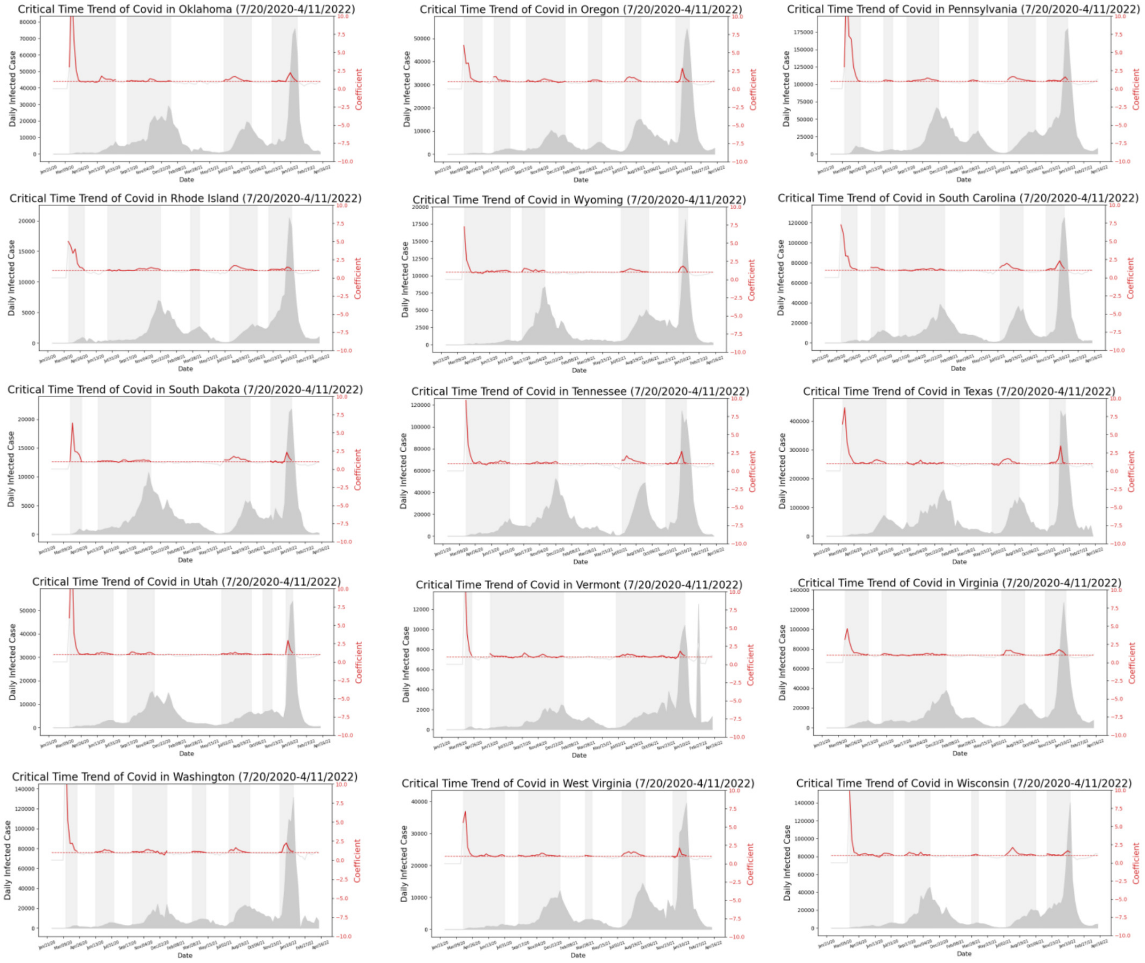


Figure S7: COVID-19 Critical Time Periods for each state in US (Part III).
